# Predicting individual clinical trajectories of depression with generative embedding

**DOI:** 10.1101/19006650

**Authors:** Stefan Frässle, Andre F. Marquand, Lianne Schmaal, Richard Dinga, Dick J. Veltman, Nic J.A. van der Wee, Marie-José van Tol, Dario Schöbi, Brenda W.J.H. Penninx, Klaas E. Stephan

## Abstract

Patients with major depressive disorder (MDD) show heterogeneous treatment response and highly variable clinical trajectories: while some patients experience swift and enduring recovery, others show relapsing-remitting or chronic disease course. Predicting individual clinical trajectories at an early disease stage is a key challenge for psychiatry and might facilitate individually tailored interventions. So far, however, reliable predictors at the single-patient level are absent.

Here, we evaluated the utility of a machine learning strategy – generative embedding – which combines an interpretable generative model with a discriminative classifier. Specifically, we used functional magnetic resonance imaging (fMRI) data of emotional face perception in 85 MDD patients from the multi-site longitudinal NEtherlands Study of Depression and Anxiety (NESDA) who had been followed up over two years and classified into three subgroups with distinct clinical trajectories. Combining a generative model of effective (directed) connectivity with support vector machines (SVMs), it was possible to predict whether a given patient will experience chronic depression vs. fast remission with a balanced accuracy of 79%. Gradual improvement vs. fast remission could still be predicted above-chance, but less convincingly, with a balanced accuracy of 61%. Importantly, generative embedding outperformed conventional (descriptive) measures such as functional connectivity or local BOLD activity, which did not predict clinical trajectories with above-chance accuracy. Furthermore, the predictive performance of generative embedding could be assigned to a specific network property: the dynamic modulation of connections by the emotional content of the trial-by-trial stimuli. Our findings suggest that a mechanistically informed generative model of a neuronal circuit underlying emotional face perception may have predictive utility for distinguishing disease courses in MDD patients.

## 1 INTRODUCTION

Major depressive disorder (MDD) is one of the most burdening mental disorders worldwide with a lifetime prevalence of 10-30% (Andrade *et al*., 2003; Kruijshaar *et al*., 2005; de Graaf *et al*., 2012). Of those patients suffering from MDD, up to a fourth are at risk of developing a chronic disease (Penninx *et al*., 2011), characterized by severe negative impact on quality of life, high rates of psychiatric comorbidities and unfavorable treatment courses (Kohler *et al*., 2019). The diagnostic criteria of MDD (DSM-5; American Psychiatric Association, 2013) are not grounded in pathophysiology, but refer to symptoms and signs (e.g., depressed mood, anhedonia, fatigue, changes in weight and sleep) that could have various causes. The diagnostic label MDD likely subsumes patients with different disease mechanisms and has limited predictive validity: patients with MDD show highly variable clinical trajectories over time (Uher *et al*., 2010; Gueorguieva *et al*., 2011; Muthen *et al*., 2011; Musliner *et al*., 2016), and the absence of mechanistically interpretable measurements or predictors turns therapy into a trial-and-error procedure (Rush *et al*., 2006; Kapur *et al*., 2012; Cuthbert and Insel, 2013). This is not only costly and frustrating for patients, but also bears the risk of long-term adverse events (McMahon and Insel, 2012) and reduced treatment adherence by the patient (Velligan *et al*., 2010).

This emphasizes the need for novel prognostic approaches to depression that furnish predictors for clinical trajectories and treatment outcomes. Predicting symptom trajectories in MDD at an early stage is of high clinical relevance because identifying patients at risk of chronic disease might guide the deployment of intensified early interventions (MacQueen, 2009). To achieve this, successful tools may benefit from being grounded in biology in order to enable a mechanistically relevant stratification of the heterogeneous MDD spectrum (Stephan *et al*., 2017). Using modern neuroimaging techniques, in particular magnetic resonance imaging (MRI) and positron emission tomography, some studies demonstrated that short-term treatment response prediction may be possible (Mayberg *et al*., 1997; Phillips *et al*., 2015; Dunlop *et al*., 2017). By contrast, it has proven more challenging to predict long-term clinical outcome, such as symptom trajectories over several years.

Schmaal et al. (2015) assessed the prognostic value of structural and functional MRI to classify disease trajectories in MDD patients from the NEtherlands Study of Depression and Anxiety (NESDA; Penninx *et al*., 2008), a multi-site longitudinal study in a large naturalistic cohort. The authors demonstrated that fMRI data from a facial emotion recognition paradigm allowed discriminating patients who, over the course of two years, showed a chronic disease trajectory from patients showing rapid remission of depressive symptoms, with up to 73% accuracy. This result was obtained by applying a supervised machine learning (ML) method (Gaussian Process Classifiers; Rasmussen and Williams, 2005) to contrast images.

While an encouraging initial result, developing this approach further with conventional ML approaches and towards clinically required levels of accuracy faces several challenges (Brodersen *et al*., 2011). First, achieving high classification accuracy robustly from whole-brain fMRI data can be difficult, given the high dimensionality of the data relative to the small sample sizes. Second, the results from “black box” ML operating on descriptive features (e.g., contrast images) do not easily allow for mechanistic interpretations. The latter, however, is increasingly recognized as critical for clinical applications of ML (e.g., Woo *et al*., 2017; Itani *et al*., 2019), both to derive novel treatment ideas from successful predictions but also to detect cases when ML goes awry, e.g., predictions that derive from artefacts in the data.

Generative embedding (GE) represents a potentially attractive alternative to “classical” ML (Shawe-Taylor and Cristianini, 2004). The idea is simple but powerful: instead of selecting features from the original data, one applies a generative model to the data and uses the ensuing model parameter estimates as features. Generative models describe how observed data may have been “generated” from latent (hidden) states or causes and thus often embody some degree of mechanistic interpretability. For example, in neuroimaging, GE uses model-based estimates of physiological or cognitive parameters, such as connection strengths (Brodersen *et al*., 2011; 2014), ion channel conductances (Symmonds *et al*., 2018), or response inhibition (Wiecki *et al*., 2016) to predict clinical states. More technically, GE views a generative model as a theory-driven dimensionality reduction device that projects high-dimensional (neuroimaging) data onto neurobiologically meaningful parameters that define a low-dimensional and interpretable space for classification. Provided one has a reasonable model, GE frequently yields more accurate results than conventional ML (e.g., Brodersen *et al*., 2011; 2014), likely because the generative model separates signal (reflecting the process of interest) from (measurement) noise.

Model-based estimates of brain connectivity might be particularly informative for predicting clinical trajectories in MDD, given that dysconnectivity has been postulated as a hallmark of depression (e.g., Mayberg, 1997; Greicius *et al*., 2007; Wang *et al*., 2012). Here, we used a generative model of fMRI data, dynamic causal modeling (DCM; Friston *et al*., 2003), to infer effective (directed) connectivity and test the utility of GE for predicting individual clinical trajectories in MDD patients from the NESDA study. For this purpose, we combined DCMs of the facial emotion recognition network with linear support vector machines (SVMs). We then compared the cross-validated predictive accuracy of GE with more conventional approaches, such as classification based on functional connectivity and local BOLD activity. We hypothesized that a biologically plausible generative model would be superior for predicting naturalistic disease courses from fMRI data.

## 2 MATERIALS AND METHODS

### 2.1 Participants

The data used in this study were acquired in the NESDA study (Penninx *et al*., 2008). NESDA is a multi-site longitudinal study to characterize the long-term course of depression and anxiety disorders in a large naturalistic cohort. In total, 2981 participants (18-65 years) were recruited from community, primary care and specialized mental health organizations. From this cohort, 301 participants (156 with MDD diagnosis) were included in the MRI experiment (for detailed descriptions of the full sample, see van Tol *et al*., 2010). For the current study, only those participants were included that had: (i) a DSM-IV diagnosis of MDD, as established using the structured Composite International Diagnostic Interview (CIDI; Robins *et al*., 1988) in the 6 months prior to baseline, (ii) reported symptoms in the month before baseline as confirmed by either the CIDI or the Life Chart Interview (LCI; Lyketsos *et al*., 1994), (iii) availability of 2-year follow-up of depressive symptoms from the LCI, and (iv) no other exclusion criteria related to, e.g., poor data quality, non-compliance with task instructions, or deficient performance (for details, see Schmaal *et al*., 2015). This resulted in a final sample of 85 participants (for an overview of the demographic and clinical characteristics, see Table 1).

**Table 1:** Demographic and clinical characteristics of participants included in the generative embedding analyses.

| Characteristic | REM (*n*=39) | IMP (*n*=31) | CHR (*n*=15) | Statistic | *p*-value |
| --- | --- | --- | --- | --- | --- |
| Age, Years | 35.90 (11.50) | 35.03 (10.00) | 44.00 (10.01) | $F=3.92$ | 0.02 |
| Gender, *n* (%) |  |  |  |  |  |
| Female | 28 (72) | 20 (65) | 9 (60) | $\chi^2=0.83$ | 0.66 |
| Male | 22 (28) | 11 (35) | 6 (40) |  |  |
| Education, Years | 11.74 (3.30) | 12.26 (3.00) | 12.00 (2.36) | $F=0.25$ | 0.78 |
| Scan Location, *n* (%) |  |  |  |  |  |
| AMC Amsterdam | 5 (13) | 5 (16) | 4 (27) | $\chi^2=3.81$ | 0.43 |
| LUMC Leiden | 13 (33) | 15 (48) | 5 (33) |  |  |
| UMCG Groningen | 21 (54) | 11 (35) | 6 (40) |  |  |
| IDS Total T1 | 31.44 (10.76) | 32.77 (8.35) | 33.72 (7.91) | $F=0.37$ | 0.69 |
| IDS Total T2 | 14.72 (9.06) | 22.29 (9.93) | 29.60 (7.17) | $F=15.87$ | < 0.001 |
| IDS Change (T2-T1) | -16.72 (11.33) | -10.48 (10.56) | -4.13 (8.10) | $F=8.35$ | < 0.001 |
| BAI Total T1 | 14.90 (9.25) | 15.77 (9.42) | 14.27 (6.93) | $F=0.16$ | 0.85 |
| Antidepressant Use T1, *n* (%) |  |  |  |  |  |
| No | 27 (69) | 22 (71) | 8 (53) | $\chi^2=1.58$ | 0.45 |
| Yes | 12 (31) | 9 (29) | 7 (47) |  |  |
| Antidepressant Use T2, *n* (%) |  |  |  |  |  |
| No | 25 (64) | 22 (71) | 9 (60) | $\chi^2=0.64$ | 0.73 |
| Yes | 14 (36) | 9 (29) | 6 (40) |  |  |

Based on the two-year follow-up clinical trajectories derived from CIDI and LCI information, MDD patients were divided into different categories with distinct courses of symptom severity. This division was informed by a latent class growth analysis (Rhebergen *et al*., 2012), as reported by Schmaal et al. (2015). The three classes were: (i) MDD-remitted, showing a rapid remission of symptoms (REM: *n*=39), (ii) MDD-improved, showing a slow but gradual improvement of symptoms from baseline to follow-up (IMP: *n*=31), and (iii) MDD-chronic, showing no improvement of symptoms from baseline to follow-up (CHR: *n*=15).

### 2.2 Experimental procedure

For fMRI, an event-related emotional face perception paradigm was used. Participants viewed color images of angry, fearful, sad, happy, and neutral facial expressions, as well as scrambled faces. Stimuli were taken from the Karolinska Directed Emotional Faces System (Lundqvist *et al*., 1998) and shown on the screen for 2.5s, with an inter-stimulus interval varying between 0.5-1.5s. Participants were instructed to indicate the gender of the presented face via button press. For scrambled images, participants had to press buttons in accordance with an arrow pointing to the left or right. Stimuli were presented using E-prime software (Psychological Software Tools, Pittsburgh, PA; https://pstnet.com/products/e-prime/). For details, see Demenescu et al. (2011).

### 2.3 Functional magnetic resonance imaging

#### 2.3.1 Image acquisition

For NESDA, structural and functional MRI data were acquired at the University Medical Center Groningen (UMCG), Amsterdam Medical Center (AMC), and Leiden University Medical Center (LUMC). Participants were scanned on 3-Tesla MR scanners (Philips Healthcare, Best, The Netherlands) with SENSE 8-channel (LUMC, UMCG) or 6-channel (AMC) receiver head coils. For details, see Supplementary Material S1.

#### 2.3.2 Image data processing

Some of the functional images were affected by a “column” or “pencil beam” artifact caused by imperfect fat suppression pulses in the Philips scanners. The artifact was most apparent in temporal signal-to-noise ratio (tSNR) maps and manifested as vertical stripes, primarily in frontal gyrus and anterior temporal lobe (see https://github.com/dinga92/stripe_cleaning_scripts). This was corrected by regressing out artifact-related independent components prior to the routine preprocessing steps. Artifact correction was done within FSL (FMRIB’s Software Library; Smith *et al*., 2004; http://www.fmrib.ox.ac.uk/fsl) as follows: First, the Multivariate Exploratory Linear Optimized Decomposition into Independent Components (*melodic*) algorithm was used to identify independent components associated with the artifact, and second, *regfilt* was used to regress out the artifact-related components.

After artifact correction, functional images were analyzed using SPM12 (Statistical Parametric Mapping, version R7487, Wellcome Centre for Human Neuroimaging, London, UK, http://www.fil.ion.ucl.ac.uk) and Matlab (Mathworks, Natick, MA, USA). Individual images were realigned to the mean image, coregistered with the high-resolution anatomical image, and normalized to the Montreal Neurological Institute (MNI) standard space using the unified segmentation-normalization approach (Ashburner and Friston, 2005). During spatial normalization, functional images were resampled to a voxel size of 2×2×2mm^3^. Finally, normalized functional images were spatially smoothed using an 8mm FWHM Gaussian kernel.

Preprocessed and artifact-corrected functional images of every participant entered first-level General Linear Model analyses (GLM; Friston *et al*., 1995) to identify brain activity related to the experimental manipulation. Each condition (i.e., angry, fearful, happy, sad, neutral, and scrambled faces) was modeled as an individual regressor, consisting of a train of stimulus onsets, which was convolved with the standard canonical hemodynamic response function (HRF). Additionally, temporal and dispersion derivatives of the canonical HRF were included to account for variability in shape and timing of hemodynamic responses (Friston *et al*., 1998). Realignment parameters were included as nuisance regressors to control for movement-related artifacts. Additionally, low-frequency fluctuations in the data were removed using a high-pass filter (cut-off 1/128Hz).

#### 2.3.3 Time series extraction

We selected six regions of interest (ROIs) that represent key components of the extended face perception network (Haxby *et al*., 2000). These ROIs were located bilaterally in the occipital face area (OFA; Puce *et al*., 1996), fusiform face area (FFA; Kanwisher *et al*., 1997), and amygdala (Breiter *et al*., 1996). To account for inter-subject variability in the exact location of these regions, center coordinates were defined for each participant individually: First, we identified the most likely MNI coordinates of these regions from a meta-analysis of 720 studies using Neurosynth (Yarkoni *et al*., 2011) with the search criterion “face”. Relying on this external information from Neurosynth helped ensure complete independence of feature selection (i.e., definition of ROI coordinates) and subsequent prediction. Generally, we prevented any cross-talk between training and test samples which might otherwise positively bias classification accuracy (see Supplementary Material S2 and Brodersen *et al*., 2011). Second, individual peak activation coordinates were defined as the subject-specific local maximum closest to the Neurosynth coordinates within a 12mm sphere. The ensuing distribution of individual coordinates is illustrated in Supplementary Figure S1. Third, BOLD signal time series were extracted from the subject-specific ROIs as the first eigenvariate of all voxels within an 8mm sphere centered around the individual coordinates. Time series were mean-centered and movement-related variance was removed (by regression using the realignment parameters).

### 2.4 Dynamic causal modeling

Dynamic causal modeling (DCM; Friston *et al*., 2003) is a generative model that enables inference on hidden (latent) neuronal states from measured neuroimaging data. For fMRI, dynamics of neuronal activity are described as a function of the effective (directed) connectivity among neuronal populations using a bilinear differential equation:

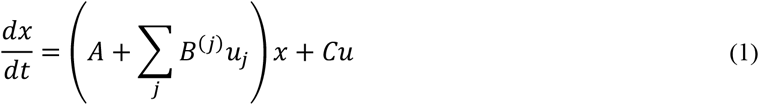

where *x* defines the neuronal states, *A* encodes the endogenous connectivity among brain regions in the absence of experimental manipulations, *B*^(*j*)^ represents the modulatory influence that input *u*_*j*_ exerts on endogenous connections, and *C* quantifies the strength of experimentally controlled inputs (perturbations) on brain regions. Integrating Eq. (1) yields a predicted neuronal time course which is then passed through a nonlinear hemodynamic model that translates neuronal signal into predicted BOLD signal (Buxton *et al*., 1998; Friston *et al*., 2000; Stephan *et al*., 2007). This yields a complete forward mapping from hidden neuronal states to observable fMRI data and, under Gaussian assumptions about the measurement noise, specifies the likelihood function. By specifying prior distributions over model parameters (neuronal, hemodynamic) and hyperparameters (measurement noise), DCM becomes a fully generative model. Model inversion then proceeds with approximate Bayesian schemes, most commonly variational Bayes under the Laplace approximation (VBL; Friston *et al*., 2007).

#### 2.4.1 Definition of model space

Inference on effective connectivity is conditional on the underlying model (e.g., assumptions about the network architecture). However, there typically exist several a priori hypotheses about the likely network structure. This model uncertainty leads to defining a model space – a set of plausible candidate models that are compared. Here, a total of seven models were constructed, representing different hypotheses of the effective connectivity in the above-mentioned network mediating perception of emotional faces. For all models, the endogenous and driving input connectivity (A- and C-matrix) were identical. Driving inputs were set to elicit face-sensitive activation (regardless of whether an emotional or neutral face was presented) in left and right OFA, consistent with their proposed role as the first stage in the face perception network (Haxby *et al*., 2000; Pitcher *et al*., 2011). The stimulus-evoked activity then propagated through the network via intra- and interhemispheric connections. We assumed forward and backward intrahemispheric connections between OFA and FFA, and between FFA and amygdala, but not between OFA and amygdala – consistent with the notion of a hierarchy in the face perception network (Haxby *et al*., 2000; Fairhall and Ishai, 2007). Additionally, reciprocal interhemispheric connections were set between homotopic regions (Zeki, 1970; Van Essen *et al*., 1982; Clarke and Miklossy, 1990; Zilles and Clarke, 1997; Catani and Thiebaut de Schotten, 2008), but omitted between heterotopic regions as these were found to be less pronounced (Hofer and Frahm, 2006; Catani and Thiebaut de Schotten, 2008).

For this basic structure, seven different modulatory connectivity patterns were defined (Figure 1), representing distinct hypotheses of how emotion processing could modulate intra- and interhemispheric connections in the extended face perception network (Fairhall and Ishai, 2007; Frässle *et al*., 2016). Emotion processing could modulate either (i) forward, (ii) backward, or (iii) forward and backward intrahemispheric connections. Similarly, emotion processing could modulate interhemispheric connections among homotopic regions or not. This yielded six different models, representing all possible combinations of the above effects. Furthermore, we included a “null” model (model 7) where none of the connections were modulated.

**Figure 1:**
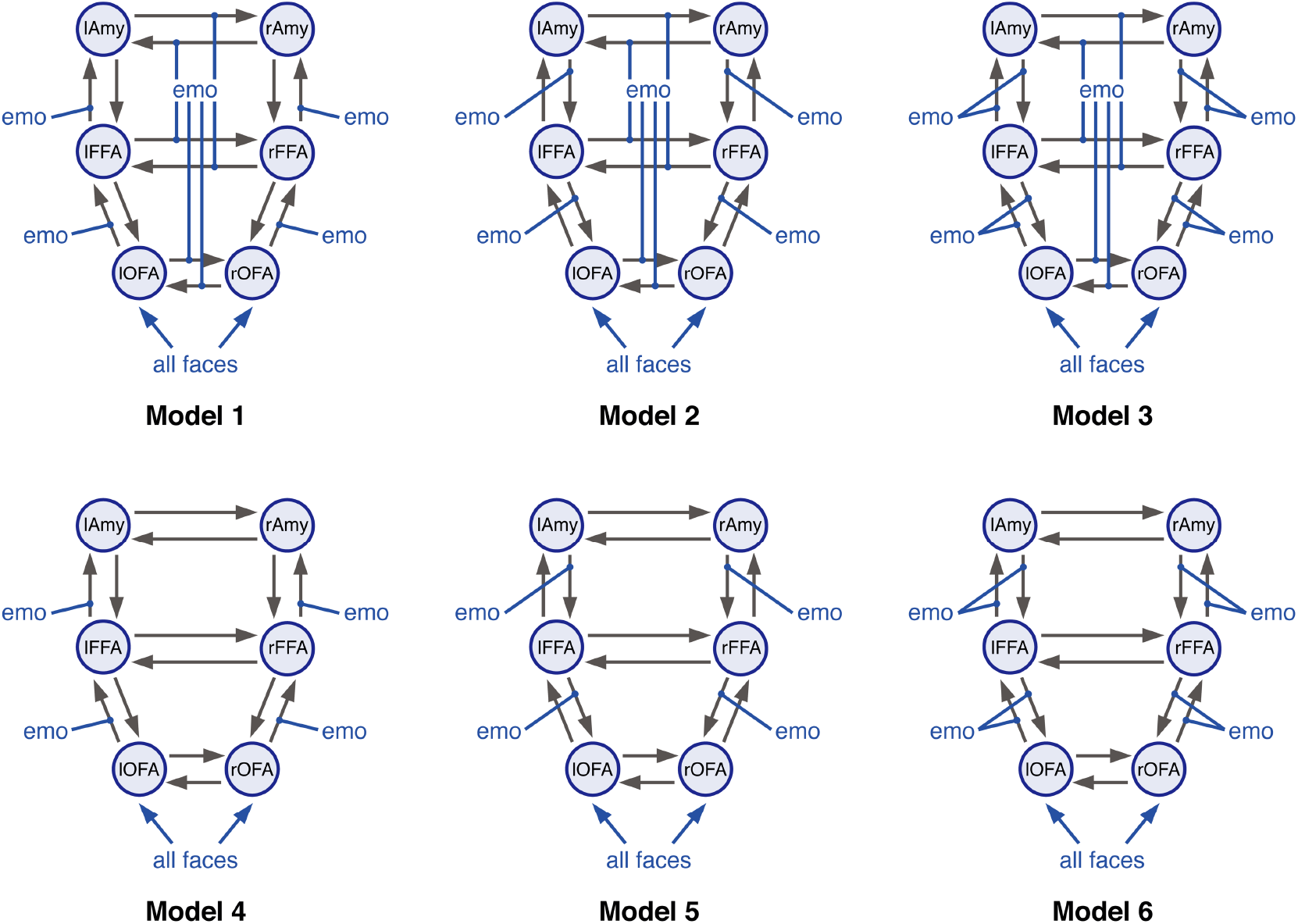
Different plausible hypotheses of the effective connectivity pattern in the network mediating emotional face perception. Forward and backward intrahemispheric endogenous connections were set between OFA and FFA, and between FFA and amygdala (Amy). Additional, reciprocal interhemispheric connections were set between bilateral OFA, bilateral FFA and bilateral amygdala. Driving inputs comprised all faces, regardless of the emotional valence, and were allowed to drive neuronal activity in the left and right OFA. While endogenous connectivity and driving inputs were identical for all models, they differed in the assumed modulatory influences of emotion processing. Emotion processing could either modulate (i) forward (models 1&4), (ii) backward (models 2&5), or (iii) forward and backward intrahemispheric connections (models 3&6). Additionally, emotion processing (i) modulated (models 1-3) or (ii) did not modulate interhemispheric connections among homotopic brain regions (models 4-6). Systematically varying all combinations resulted in six distinct models. Finally, we also included a “null” model (i.e., model 7, not shown) where none of the intra- and interhemispheric connections was modulate by emotion processing.

Driving and modulatory inputs were not mean-centered. Model inversion was performed using DCM12 (SPM12, version R7487). For details, see Supplementary Material S3.

#### 2.4.2 Bayesian model averaging

We computed individual parameter estimates by means of Bayesian model averaging (BMA; Penny *et al*., 2010) across all models in our model space within a pre-specified Occam’s window (*π*_*occ*_ = 0.05). For details, see Supplementary Material S4. BMA parameter estimates represent a weighted average across the considered models, where each model contributes according to its posterior model probability. Importantly, in order to prevent any potential cross-talk between training and test sample, BMA parameters were computed for each participant individually.

### 2.5 Generative embedding

The ensuing means of the posterior densities of BMA parameters from each participant were used to create a generative score space for a discriminative classification method. In total, this yielded 78 parameters/features. Within this space, a linear kernel representing the inner product *k(x*_*i*_, *x*_*j*_ ) = ⟨*x*_*i*_, *x*_*j*_⟩ was used to compare two instances (participants). A support vector machine (SVM) was then applied for binary classification of pairwise combinations of the three MDD groups (i.e., REM, IMP, and CHR). Specifically, we used the *fitcsvm* routine in Matlab. An estimate of classification performance was obtained by means of a leave-one-out cross-validation procedure. Here, in each fold, the classifier is trained on *n* − 1 participants (the training set) and tested on the left-out participant. Using the training set only, the hyperparameters of the SVM (i.e., box constraint and kernel scale; see Supplementary Material S5) were optimized using in-built routines of *fitcsvm*. This computes the Bayes-optimal hyperparameters using the expected improvement acquisition function (Frazier, 2018) based on an (inner) five-fold cross validation. This approach is known as nested cross-validation (Stone, 1974; Cawley and Talbot, 2010). By default, *fitcsvm* solves SVMs using the Sequential Minimal Optimization algorithm (SMO; Fan *et al*., 2005). Significance of the classification result was assessed by means of permutation tests. Here, an empirical null distribution of the balanced accuracy is computed by randomly permuting the participant labels and re-fitting the entire classification model (i.e., training and testing) based on these new labels (Good, 2000; Ojala and Garriga, 2010). For each permutation, the balanced accuracy is re-evaluated. Here, we used 1,000 permutations. The *p*-value is then computed as the rank of the original balanced accuracy in the distribution of permutation-based balanced accuracies, divided by the total number of permutations.

### 2.6 Data availability

The data utilized in this study are available as part of the NESDA consortium upon request.

## 3 RESULTS

### 3.1 Classification of clinical trajectories

#### 3.1.1 Predictive accuracy of effective connectivity parameters

Our results suggest that DCM parameter estimates discriminated patients with a chronic disease trajectory from patients that show fast remission with a balanced accuracy of 79% (*p*<0.001; Figure 2, *blue*). We then evaluated the underlying receiver-operating characteristic (ROC) and precision-recall (PR) curves (Figure 3A+B). From the ROC curve, the respective area under the curve (AUC) for discriminating chronic from remitting patients evaluated to 0.87 (*blue curve*). Notably, high recall (sensitivity) might come at the expense of low positive predictive value (PPV; also known as “precision”) – particularly, in the presence of class imbalances. This is problematic for clinical applications where one strives to maximize sensitivity while at the same time keeping PPV uncompromised. In reality, there is always a trade-off between the two. Hence, we also inspected the PR curves and found that our classifier achieved 97% sensitivity at a PPV of 86% when discriminating chronic from remitting patients.

**Figure 2:**
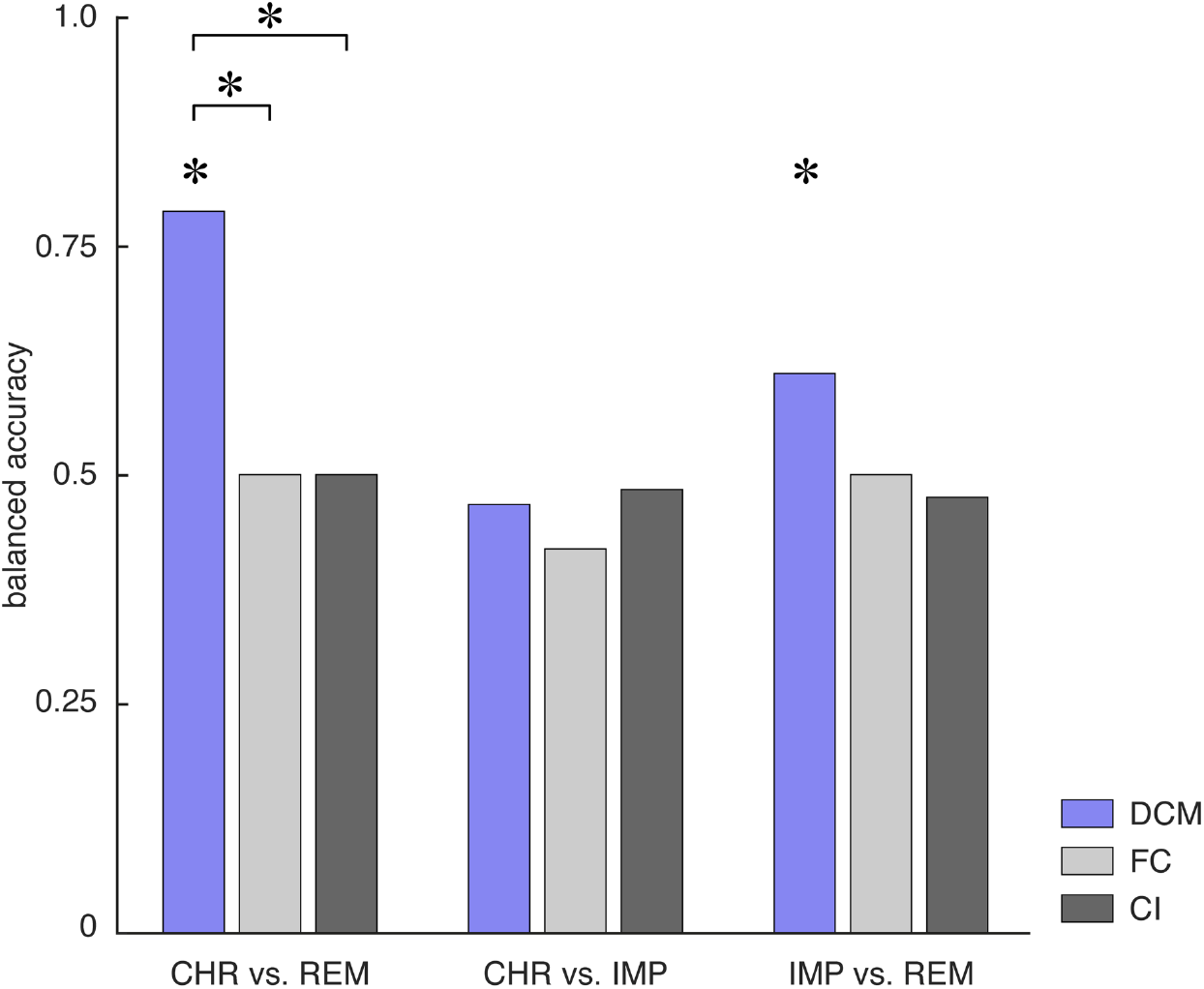
Balanced accuracy for the binary classifiers as assessed using leave-one-out cross validation for the three different subgroup comparisons – that is, CHR vs. REM (*left*), CHR vs. IMP (*middle*), and IMP vs. REM (*right*). Balanced accuracies are shown for the different features – namely, effective connectivity parameters (DCM; *blue*), functional connectivity (FC; *light grey*), and local BOLD activity (CI, *dark grey*). Asterisks above the bars indicate significant classification performance as assessed by means of permutation tests where an empirical null distribution of the balanced accuracy is computed by randomly permuting the participant labels and re-evaluating the classifier based on these new labels. Additionally, asterisks above the lines connecting two bars indicate significant differences in classification performance between different data features as assessed using the asymptotic McNemar test.

**Figure 3:**
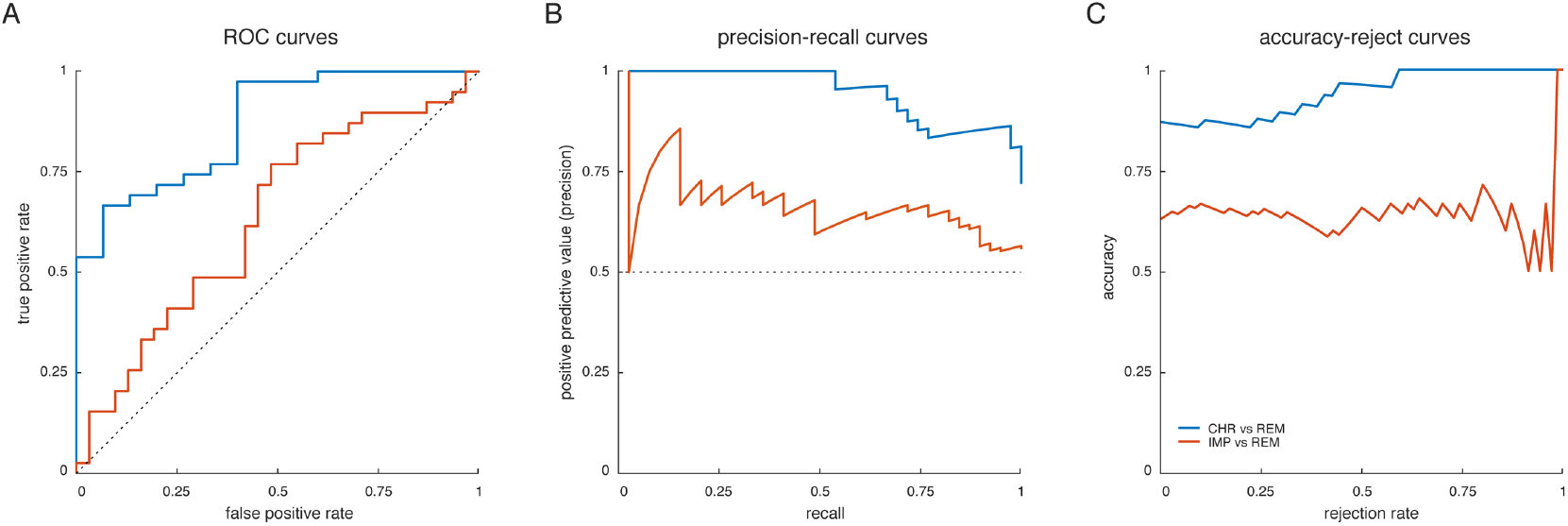
Performance curves for the two binary classifiers that achieved above-chance balanced accuracies – that is, CHR vs. REM (*blue curve*) and IMP vs. REM (*red curve*). **(A)** receiver-operating characteristic (ROC) curves, illustrating the trade-off between the true positive rate (sensitivity) and the false positive rate (1-specificity) across the entire range of detection thresholds, **(B)** precision-recall (PR) curves, illustrating the trade-off between the precision (positive predictive value) and recall (true positive rate) for different thresholds, and **(C)** accuracy-reject curves, representing the accuracy of a classifier as a function of the rejection rate (Nadeem *et al*., 2010). For a comprehensive summary of all classification results, see Table 2.

Effective connectivity parameters further discriminated between patients that showed gradual improvement of depressive symptoms and patients with fast remission with a balanced accuracy of 61% (*p*=0.03; Figure 2), although this did not reach significance when correcting for multiple comparisons (*α*_*Bonf*_=0.0056). The AUC was 0.63 (Figure 3A+B; *red curve*). In contrast, chronic patients could not be differentiated from those with gradual improvement above-chance level (balanced accuracy: 47%, *p*=0.92; Figure 2), corresponding to an AUC of 0.35. Table 2 provides a comprehensive summary of all classification results.

**Table 2:** Classification results for the generative embedding procedure. Shown are key performance measures of the classification algorithm, including: balanced accuracy, area under the curve, sensitivity (recall), specificity, positive predictive value (precision), and negative predictive value. Performance measures are shown for the three different binary classifications (i.e., CHR vs. REM, CHR vs. IMP, and IMP vs. REM).

| Classification | CHR ($n=15$)<br>vs. REM ($n=39$) | CHR ($n=15$)<br>vs. IMP ($n=31$) | IMP ($n=31$) vs.<br>REM ($n=39$) |
| --- | --- | --- | --- |
| Accuracy | 0.87 | 0.63 | 0.63 |
| Balanced accuracy | 0.79 | 0.47 | 0.61 |
| Area under the curve (AUC) | 0.87 | 0.35 | 0.63 |
| Sensitivity (recall) | 0.97 | 0.94 | 0.77 |
| Specificity | 0.60 | 0 | 0.45 |
| Positive predictive value (Precision) | 0.86 | 0.66 | 0.64 |
| Negative predictive value | 0.90 | 0 | 0.61 |

Since groups differed significantly in age (but no other variable; Table 1), we repeated the analysis after regressing out age as a confound from the DCM parameters. We found results to be highly consistent (although with slightly decreased accuracies), suggesting that results are not simply caused by age (Supplementary Material S8).

#### 3.1.2 Comparison to functional connectivity and fMRI activity

We compared the GE results to an alternative approach in which the classifier operates on estimates of functional connectivity. Following standard practice, functional connectivity was computed in terms of Pearson’s correlations among the same BOLD signal time series that had previously been used for the DCMs. However, functional connectivity measures did not discriminate between the different clinical trajectories above chance, with balanced accuracies of 50% (*p*=0.77) for CHR vs. REM patients, 42% (*p*=0.996) for CHR vs. IMP patients, and 50% (*p*=0.37) for IMP vs. REM patients (Figure 2, *light grey*). Importantly, for discriminating chronic from fast remitting patients, GE significantly outperformed functional connectivity measures (*p*=0.01; asymptotic McNemar test^1^ as implemented in MATLAB’s *testcholdout* function).

In addition, we tested whether the different clinical trajectories could be distinguished based on measures of local BOLD activity within the same regions utilized above. Local BOLD activity was quantified in terms of the regional mean and standard deviation of the contrast estimates within the 8mm spheres for all face-related contrasts (i.e., contrasts representing the individual regressors in the first-level GLM encoding angry, fearful, happy, sad, and neutral faces; see Methods). Again, the different clinical trajectories were indistinguishable based on mere estimates of local BOLD activity, with balanced accuracies of 50% (*p*=0.72) for CHR vs. REM patients, 48% (*p*=0.81) for CHR vs. IMP patients, and 48% (*p*=0.51) for IMP vs. REM patients (Figure 2, *dark grey*). As for functional connectivity, GE significantly outperformed local BOLD activity for distinguishing chronic patients from those that showed fast remission, (*p*=0.01).

### 3.2 Assessment of predictive confidence

Next, we computed accuracy-reject curves for the two binary classifiers that achieved above-chance balanced accuracies (i.e., CHR vs. REM, IMP vs. REM) to assess the predictive confidence of our GE approach. Accuracy-reject curves demonstrate the accuracy of a classifier when only predictions greater than a certain (relative) confidence threshold are considered (Nadeem *et al*., 2010). Hence, this resembles classification with a reject option (Bishop, 2006), where cases that do not meet a certain confidence criterion can be deferred to a clinician for further inspection. We found that for distinguishing chronic patients from those with fast remission, the classifier yielded perfect classification accuracy at a rejection threshold of 60% of participants (Figure 3C; *blue curve*). Furthermore, the accuracy-reject curve overall increased as function of the rejection rate, suggesting that participants that lie further from the decision hyperplane were more likely to be assigned correctly to their respective class. In contrast, for distinguishing gradually improving patients from those showing fast remission no such cut-off could be identified and the curve did not reveal a steady increase as a function of the rejection rate (Figure 3C; *red curve*).

### 3.3 Inspection of the generative score space

One benefit of GE is that features represent model parameter estimates, which, depending on the model, may be neurobiologically interpretable. Hence, in a next step, we interrogated our generative score space to illustrate which features contributed most to the classification performance.

#### 3.3.1 Inspection of individual predictive features

In a first step, we aimed to pinpoint the individual contribution of each feature (i.e., DCM parameter) separately for the two significant classifiers (i.e., CHR vs. REM, IMP vs. REM). Importantly, individual feature weights of linear classifiers are not directly interpretable because high magnitudes of feature weights might indicate an association with the label or a “suppressor” variable that cancels out noise or mismatch in other colinear variables (Naselaris *et al*., 2011; Haufe *et al*., 2014). Therefore, we followed the procedure described by Haufe *et al*. (2014) and first transformed all feature weights into patterns based on a corresponding forward mapping.

For both classifiers, the features that received the highest average (across cross-validation folds) scores were situated along the dimension of modulatory (emotional) influences (Figure 4, *top*), whereas the endogenous connectivity and driving input parameters did not score highly and thus did not distinguish strongly between the groups. Importantly, since averaging over cross-validation folds might artificially smooth the weights due to correlations among folds, we inspected the variability of the observed results across the individual cross-validation folds. This suggested that the observed pattern was highly consistent for both classifiers (Figure 4, *bottom*).

**Figure 4:**
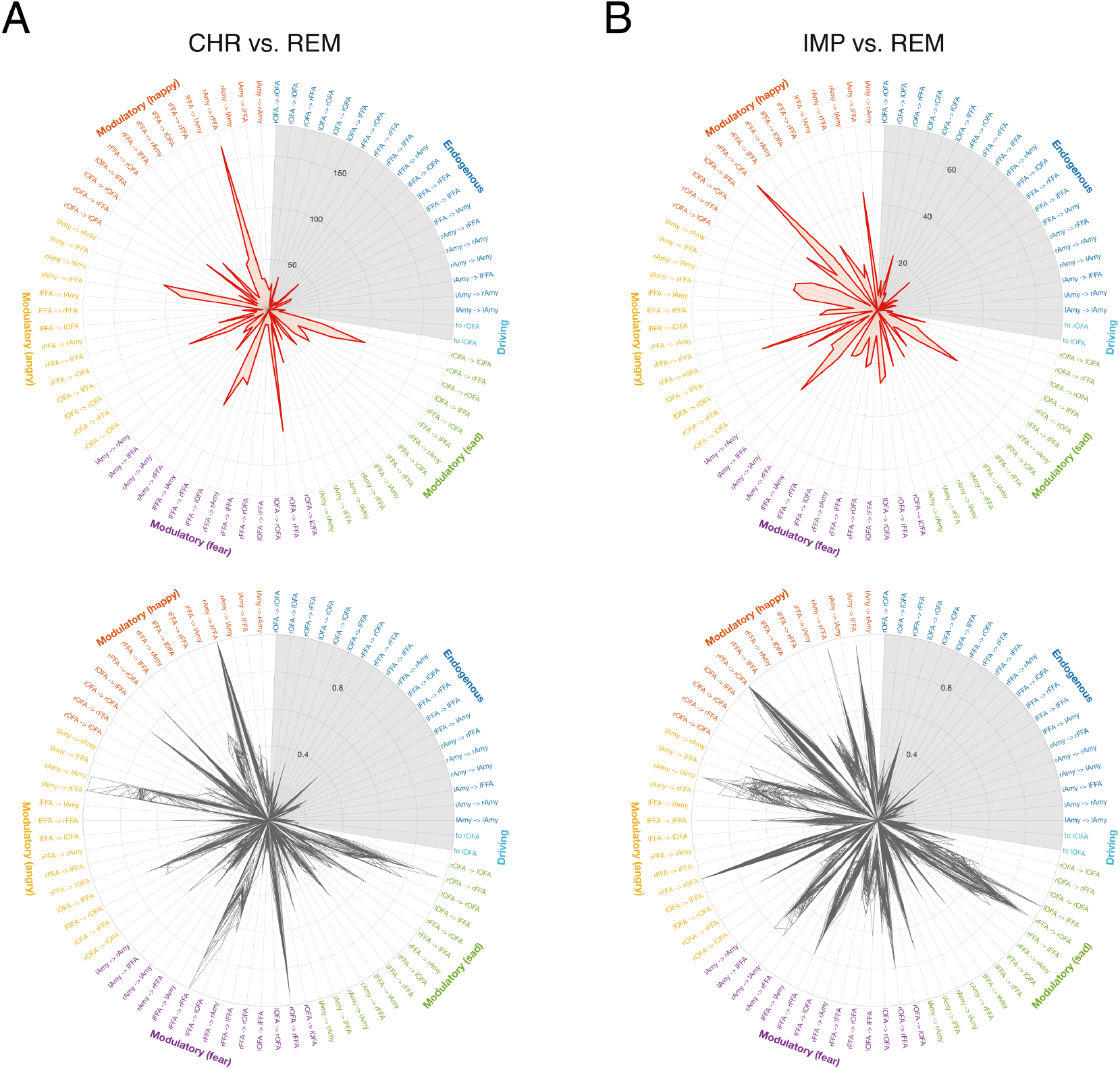
Illustration of the relevance of individual features. First, feature weights were transformed into feature patterns to allow for interpretability (Haufe *et al*., 2014). The respective score of each individual feature (DCM parameter) is then shown as a polar plot for the classifier distinguishing **(A)** patients with a chronic disease trajectory from patients that showed fast remission (CHR vs. REM), and **(B)** patients with gradual improvement of symptom severity from patients that showed fast remission (IMP vs. REM). (*Top*) Magnitude of scores computed as the average across all cross-validation folds, (*bottom*) magnitude of scores for each cross-validation fold individually, normalized to the maximum score within each fold for displaying purposes. The grey area represents endogenous connectivity and driving input parameters, showing less pronounced scores as the modulatory parameters. Endogenous connectivity is colored in blue, modulatory influences of happy faces in red, modulatory influences of angry faces in yellow, modulatory influences of fearful faces in violet, modulatory influences of sad faces in green, and driving inputs (related to all faces regardless of the emotional valence) in cyan.

In brief, for distinguishing chronic from fast remission patients, the modulatory influence of happy faces on the connection from right amygdala to right FFA received the highest score (Figure 4A). Furthermore, scores were high for modulatory influences of negative emotions (i.e., fear, anger, and sadness) on the functional integration among face-processing regions and emotion-sensitive regions. For instance, modulatory influences by angry faces on the connection from right amygdala to right FFA and left amygdala, and on the connection from left OFA to left FFA showed high loads. Similarly, the modulation of connections from right OFA and left FFA to right FFA, as well as the connection from right FFA to right amygdala by fearful faces received high scores.

For distinguishing gradually improving patients from those that showed fast remission, the modulatory influence of happy faces on the connection from right FFA to right OFA received the highest score (Figure 4B). Modulatory influences by angry and sad faces on this connection also showed high loads. Similarly, modulation of connections among FFA and amygdala in both hemispheres by angry faces scored highly, as well as modulation of various endogenous connections (e.g., left amygdala to left FFA, left to right OFA, left FFA to left OFA) by happy faces.

In summary, chronic and gradually improving patients could be differentiated from patients with a benign clinical trajectory (i.e., fast remission) primarily in terms of how emotions modulated the functional integration within the extended face perception network.

## 4 DISCUSSION

The analyses presented in this paper demonstrate the utility of GE for predicting individual clinical trajectories of MDD patients over a two-year period. Using fMRI data from the NESDA study and DCM to infer effective connectivity within the facial emotion perception network, model parameter estimates served as features for supervised learning. This GE approach enabled the prediction of whether a given patient would show a chronic disease course or fast remission, with a balanced accuracy of 79%. Additionally, patients with gradual improvement in symptom severity could be distinguished from those who remitted quickly with a balanced accuracy of 61%. Notably, GE outperformed more conventional (descriptive) measures, i.e., functional connectivity or local BOLD activity within the network of interest. Similar to previous studies (Brodersen *et al*., 2011; 2014), these findings demonstrate that using a plausible generative model as the basis for classification can enhance classification accuracy significantly.

Apart from the superior classification accuracy, another advantage of GE is that results can be interpreted in terms of the mechanisms represented by the underlying generative model. To this end, one can interrogate the generative score space to identify the (sets of) features that are most discriminative between the different classes (Brodersen *et al*., 2011). Here, we addressed this by first transforming the feature weights of the linear SVM into patterns, following previous recommendations (Haufe *et al*., 2014). Inspecting these scores then allowed to pinpoint the features that contributed most strongly to the classification between the different naturalistic courses (Figure 4). This analysis suggested that groups differed primarily along the dimension encoded by the modulatory parameters, which represent changes in the endogenous connectivity by emotional valence of the face stimulus presented. Put differently, it is the dynamic (trial-by-trial) modulation of connections by the emotional contents of the faces that allows for predicting which clinical trajectory an individual patient will experience – not the average connectivity across all trials. In conclusion, our analysis implies that it is the reactivity of the face processing network to emotional stimuli – in terms of reconfiguring its connection strengths in a trial-wise fashion – which enables prediction of the future clinical trajectory of an individual patient.

These results are consistent with our findings from group comparisons of the connectivity patterns (see Supplementary Material S7), which, however, do not allow for single-subject predictions. Our results are also consistent with previous work suggesting aberrant processing and regulation of emotions as a key pathology underlying MDD (Harmer *et al*., 2009; Rive *et al*., 2013). For instance, an fMRI meta-analysis demonstrated valence-dependent effects of emotional stimuli on amygdala and fusiform gyrus in depression, with hyperactivation for negative and hypoactivation for positive stimuli (Groenewold *et al*., 2013; but see Muller *et al*., 2017). Along these lines, reduced amygdala activity to positive emotional stimuli has been associated with anhedonia (Stuhrmann *et al*., 2013). Similarly, functional integration among key components of the emotion processing network has been found to be altered in MDD (Mayberg, 1997; Greicius *et al*., 2007; Almeida *et al*., 2009). Alterations in emotion processing have also been suggested to have some clinical utility. For instance, implicit processing of affective facial expressions predicted a diagnosis of MDD (Fu *et al*., 2007), and longitudinal neuroimaging studies have reported normalization of aberrant activation in the neuronal circuit supporting emotion processing in response to pharmacotherapy, in particular SSRIs (Sheline *et al*., 2001; Fu *et al*., 2004; Anand *et al*., 2007; Robertson *et al*., 2007; Murphy *et al*., 2009; Godlewska *et al*., 2012; Ai *et al*., 2019).

The classification accuracies we found are comparable to the results obtained by Schmaal et al. (2015). For chronic versus swiftly remitting patients, our GE procedure yielded a higher predictive accuracy than the best result reported by Schmaal and colleagues (balanced accuracy: 73%) when not accounting for age. For the other two classifiers, our procedure yielded somewhat complementary results to Schmaal et al. (2015): While the previous work could distinguish between chronic and gradually improving patients but not between gradually improving and fast remitting patients, the opposite held for GE. This may be due to differences in classification procedure and features: Schmaal et al. (2015) used Gaussian Process Classifiers (Rasmussen and Williams, 2005) on whole-brain contrast images, whereas the present study applied linear SVMs to DCM parameter estimates from a small (six-region) network. Furthermore, their analysis was still based on the artifact-confounded MR data (see Methods); hence, it remains to be tested whether classification accuracy would change when the artifact-corrected images are used.

Previous attempts to obtain single-patient predictions in MDD have almost exclusively concerned short-term treatment responses to specific interventions. For instance, pioneering PET studies demonstrated that cingulate metabolism predicts treatment responses (Mayberg *et al*., 1997). For fMRI, Fu *et al*. (2008) showed that brain activity during the processing of sad facial expressions allows predicting treatment outcome to antidepressant medication in individual patients with major depression. Graph-theoretical measures based on functional connectivity in the default mode network at baseline were predictive of changes in symptom severity after two weeks of medication (Shen *et al*., 2015). Furthermore, activation (Siegle *et al*., 2012) and functional connectivity measures (Crowther *et al*., 2015; Walsh *et al*., 2017) were predictive of psychotherapy outcome. Similarly, functional connectivity of the subcallosal cingulate cortex with insula, dorsal midbrain and ventromedial prefrontal cortex was differentially associated with remission and treatment failure to cognitive-behavioral therapy and antidepressant medication (Dunlop *et al*., 2017). Finally, Nord *et al*. (2019) showed that neural activity at baseline was predictive of clinical responses to transcranial direct current stimulation (tDCS) to left prefrontal cortex in unmedicated MDD patients.

Arguably, the attempt to predict outcome after two years in a naturalistic setting, as in NESDA, represents a greater challenge than predicting short-term response to a particular treatment. NESDA (1) recruited patients from a wide spectrum, including community, primary care and specialized mental health organizations, (2) encompassed a wide range of depressive phenotypes from very mild to severe, and (3) did not standardize treatments or occurrence of life events over the 2-year follow-up period (Penninx *et al*., 2008). This represents a strength of the NESDA dataset since it allows testing the course of MDD in a naturalistic and realistic setting where the patient sample reflects the clinical heterogeneity that physicians face on a day-by-day basis.

Existing attempts to predict MDD trajectories have focused on clinical or cognitive features (Vreeburg *et al*., 2013; Vogelzangs *et al*., 2014; Kessler *et al*., 2016). Recently, Dinga *et al*. (2018) systematically assessed the predictive value of non-imaging data, using clinical, psychological and biological measures from the NESDA study. They found that clinical measures performed best with balanced accuracies around 66%, while endocrine and immunological measures (e.g., cortisol, inflammatory markers, metabolic syndrome markers) did not distinguish between clinical trajectories. Interestingly, consistent with our GE results, Dinga and colleagues could primarily discriminate remitted patients from the other two groups.

Few studies also attempted to predict the onset of MDD. For instance, Pan *et al*. (2017) showed that aberrant functional connectivity of the ventral striatum was predictive of the onset of depression in a community sample of adolescents. Similarly, functional connectivity during the resting state predicted onset of MDD (Hirshfeld-Becker *et al*., 2019).

Our study is subject to several limitations. First, while our sample size (*n*=85) does not fare badly compared to other imaging-based prediction studies on MDD, the sample size is still modest for attempts to obtain individual predictions, and we do not have access to a separate validation set at the present time. Second, the classical DCM approach employed here is restricted to small networks that comprise only a small number of regions to keep model inversion computationally feasible (Daunizeau *et al*., 2011; Frässle *et al*., 2018b). Consequently, we focused on a six-region network comprising only core regions of the extended face perception network (i.e., OFA, FFA and amygdala; see Haxby *et al*., 2000). However, depression is often characterized in terms of widely distributed alterations in network organization (Mayberg, 1997; Greicius *et al*., 2007; Wang *et al*., 2012) and, hence, inferring effective connectivity in MDD patients at the whole-brain level might be an appealing next step. This could be achieved by exploiting recent advances in generative models that are computationally extremely efficient and can provide whole-brain estimates of effective connectivity (Frässle *et al*., 2017; 2018a).

Another potential limitation is the two-step procedure of GE where the inversion of the generative model is independent of the subsequent ML step. An alternative is to construct a fully hierarchical generative model that describes individual data generation and simultaneously assigns participants to clusters. Such a model has recently been introduced to neuroimaging as “hierarchical unsupervised generative embedding” (HUGE; Raman *et al*., 2016; Yao *et al*., 2018). This framework might be beneficial for predicting distinct disease courses because model inversion at the single-subject level can be informed by group-level results and, vice versa, the formation of clusters takes into account the full posterior distribution of single-subject estimates.

Furthermore, in line with Schmaal *et al*. (2015), we used binary classifiers which can only distinguish between two disease trajectories. However, this approach does not allow for single class predictions, which would rest on multi-class classification (Bishop, 2006). Extending our classification scheme beyond binary classification is likely to be of clinical relevance, as multi-class prediction more faithfully resembles the decision process that physicians routinely engage in.

Finally, on a more general note, any biomarker in psychiatry will always yield imperfect predictions. This is because the course of psychiatric disorders is affected by a plethora of environmental factors, which cannot be foreseen from physiological data; e.g., the occurrence of stressful life events like loss, bereavement, or trauma (Horesh *et al*., 2008). Such external perturbations likely upper-bound the predictive accuracy of any biomarker, whether derived from neuroimaging or genetics.

Despite these limitations, the present study demonstrates the potential of GE for addressing important clinical problems in psychiatry in a way that combines enhanced accuracy with biological interpretability of predictions. More generally, as illustrated by recent successful clinical applications (e.g., Symmonds *et al*., 2018), generative models offer an attractive strategy for establishing computational assays that could inform clinical decision-making in psychiatry (for review, Frässle *et al*., 2018b). A critical condition for the future success (or failure) of this strategy will be the availability of large prospective patient datasets that, like NESDA, offer clinically relevant outcome data and allow for testing the generalizability and robustness of model-based clinical predictions in real-world settings.

## Data Availability

The data utilized in this study are available as part of the NESDA consortium upon reasonable request.

## 5 ACKNOWLEDGEMENTS

The authors would like to thank all NESDA participants for taking part in the study. Furthermore, we would like to thank Jakob Heinzle for valuable advice.

## 6 FUNDING

This work was supported by the ETH Zurich Postdoctoral Fellowship Program (SF), the Marie Curie Actions for People COFUND Program (SF), and the University of Zurich Forschungskredit Postdoc (SF), by a NHMRC Career Development Fellowship (1140764; LS), as well as by the René and Susanne Braginsky Foundation (KES) and the University of Zurich (KES). Furthermore, the infrastructure for the NESDA study (www.nesda.nl) is funded through the Geestkracht program of the Netherlands Organisation for Health Research and Development (ZonMw, grant number 10-000-1002) and financial contributions by participating universities and mental health care organizations (VU University Medical Center, GGZ inGeest, Leiden University Medical Center, Leiden University, GGZ Rivierduinen, University Medical Center Groningen, University of Groningen, Lentis, GGZ Friesland, GGZ Drenthe, Rob Giel Onderzoekscentrum).

## 7 COMPETING INTERESTS

The authors report no competing interests.

## Supplementary Material

### Supplementary Material S1: Image acquisition

For NESDA, data were acquired at the University Medical Center Groningen (UMCG), Amsterdam Medical Center (AMC), and Leiden University Medical Center (LUMC). Participants were scanned on 3-Tesla MR scanners (Philips Intera, Best, The Netherlands) with SENSE 8-channel (LUMC, UMCG) or SENSE 6-channel (AMC) receiver head coils. A T_2_^*^-weighted gradient-echo echo-planar imaging sequence was used to provide blood oxygen level dependent (BOLD) contrast. The scanning parameters differed slightly across sites: UMCG (39 slices, TR=2300ms, TE=28ms, matrix size 64×64 voxels, voxel size 3×3×3mm^3^, slice gap 0mm, flip angle 90°), AMC (35 slices, TR=2300ms, TE=30ms, matrix size 96×96 voxels, voxel size 2.29×2.29×3mm^3^, slice gap 0mm, flip angle 90°), and LUMC (35 slices, TR=2300ms, TE=30ms, matrix size 96×96 voxels, voxel size 2.29×2.29×3mm^3^, slice gap 0mm, flip angle 90°). Slices were acquired with an interleaved acquisition schedule parallel to the intercommissural (AC-PC) plane. Additionally, a high-resolution anatomical image was acquired for each participant using a T1-weighted imaging sequence (170 slices, TR=9ms, TE=3.5ms, matrix size 256×256, voxel size 1×1×1mm^3^)

### Supplementary Material S2: Unbiasedness of generative embedding procedure

When performing classification, it is critical to avoid any optimistic bias in assessing the classification performance (Brodersen *et al*., 2011). To this end, a classifier must be applied to test data that have not been used during training. In the case of generative embedding (GE), this implies that the specification of the generative model (e.g., the definition of the regions of interest) cannot be treated in isolation from its use for classification. In the present paper, we prevented any cross-talk between training and test samples using the strategy outlined below (for a graphical illustration of our workflow, see Supplementary Figure S2):

First, preprocessing of the functional magnetic resonance imaging data as well as first-level General Linear Model (GLM; Friston *et al*., 1995) analysis were performed for each participant individually, thus, avoiding any potential cross-talk between training and test samples at this stage.

Second, we ensured that the individual center coordinates of the regions of interest (ROIs) were unbiased (cf. Figure 2, Brodersen *et al*., 2011). The typical strategy for defining individual center coordinates would be to search for the nearest maximum from the group-level maximum. However, this approach is problematic since the group-level maximum would be based on data from all participants, including the left-out test participant in a subsequent leave-one-out cross-validation scheme. Consequently, training and testing samples would no longer be independent; thus, violating the fundamental assumption of cross-validation. This scenario is known as peeking (Pereira *et al*., 2009). To avoid this scenario, we used external (to the study) information from Neurosynth (Yarkoni *et al*., 2011). Specifically, we identified the most likely MNI coordinates from a meta-analysis of 720 studies using Neurosynth with the search criterion “face”. Individual peak activation coordinates were then defined as the subject-specific local maximum closest to the Neurosynth coordinates within a 12mm sphere.

Third, model inversion of each DCM is performed for each participant separately, thus, avoiding any potential cross-talk between training and test samples at this stage.

Fourth, parameter estimates from all DCMs are averaged using Bayesian model averaging (BMA; Penny *et al*., 2010). Again, the conventional strategy of performing random-effects BMA to infer group-level and single-subject BMA parameter estimates would result in the above-mentioned phenomenon of peeking and thus to positive bias in the classification performance. To avoid this scenario, BMA parameter estimates were computed for each participant individually, thus, preventing any bias. The ensuing DCM parameter estimates can then be safely used in subsequent classification procedures.

### Supplementary Material S3: Model inversion of DCMs

Model inversion (inference) of DCMs proceeded in a fully Bayesian framework using the default VBL scheme in SPM (Friston *et al*., 2007). This approach yields both the posterior distribution over model parameters and hyperparameters, as well as an estimate of the negative free energy which serves as a lower bound approximation of the log model evidence (i.e., the probability of the data given the model). Notably, VBL rests on a gradient-based optimization of the negative free energy and is therefore susceptible to local extrema in the objective function (Daunizeau *et al*., 2011). This can lead to suboptimal inference on both parameters and model evidence. Indeed, the effect of local extrema became apparent in our analysis in the form of some “flat-lined” DCMs for which the VBL algorithm converged almost immediately without notable deviations of the posterior from the prior mean. To address this issue, we performed a multi-start approach where inversion of each model in every participant was performed for 100 random starting values (not to be confused with the priors). In brief, we sampled 99 starting values from the prior density. One additional starting value corresponded to the default starting value of the gradient ascent in SPM12 (i.e., the prior mean of the parameters). The same starting values were used for all participants and models tested. Notably, model parameters representing modulatory influences were not included in the multi-start procedure, but started always from the prior means. This was necessary to ensure identical starting values across the seven models (which differed in their modulatory connectivity patterns). The best solution for each participant and model was then chosen as the one that maximized the negative free energy.

### Supplementary Material S4: Bayesian model averaging

We computed individual parameter estimates by means of Bayesian model averaging (BMA; Penny *et al*., 2010). BMA allows obtaining model parameter estimates that represent a weighted average across the considered models, thus taking into account model uncertainty. Specifically, each model contributes to this average according to its posterior model probability. For instance, the BMA posterior for participant *n*, marginalized over all models *m* of the model space *f*:

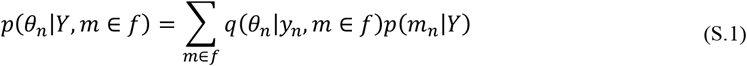

where *q*(*θ*_*n*_|*y*_*n*_, *m* ∈ *f*) ≈ *p*(*θ*_*n*_ | *y*_*n*_, *m* ∈ *f*) is the Variational approximation *q*(·) to the true subject-specific posterior density *p*(·). Furthermore, *p*(*m*_*n*_|*Y*) is the posterior model probability. More technically, BMA allows to marginalize over models – that is, average out any dependency of the parameter estimates on the assumptions about any particular model. BMA provides access to the full posterior density on model parameters, from which MAP estimates can be harvested for subsequent machine learning steps (e.g., classification).

In line with standard procedures, BMA parameter estimates in the present paper were computed by averaging across all models in our model space within SPM12’s default Occam’s window (*π*_*occ*_ = 0.05). This means that models with a low posterior model probability in relation to the most likely model are excluded from the BMA estimation (Madigan and Raftery, 1994). This Occam’s window is chosen for computational expedience only. Importantly, in Eq. S.1, the BMA posterior depends on the entire dataset *Y* = *y*_*1*…*N*_, including the left-out test participant in a subsequent leave-one-out cross-validation scheme. In order to prevent this cross-talk between training and test sample, BMA parameters were computed for each participant individually:

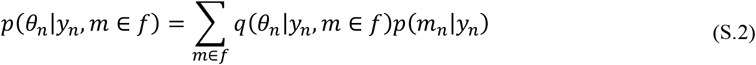

yielding a BMA posterior that depends only on the data *y*_*n*_ from the respective participant (see Supplementary Figure S2).

### Supplementary Material S5: Support vector machine implementation in *fitcsvm*

In the present paper, binary classification was performed using a linear support vector machine (SVM) as implemented in the *fitcsvm* routine in Matlab. For the non-separable case (i.e., whenever the training data is not perfectly separable), also called soft-margin SVM, the mathematical formulation of the SVM takes the form:

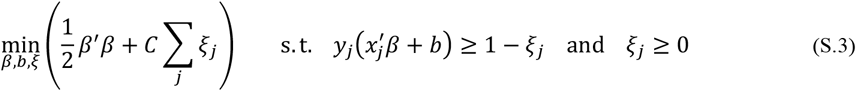

where *β* is the weight vector *b* is the bias, and *ξ*_*j*_ is the *j*-th slack variable, which causes a penalty weighted by the parameter *C*. Hence, *C* controls the maximum penalty imposed on margin-violating observations and larger values lead to a stricter separation. In the machine learning literature, *C* is typically referred to as the “cost” parameter of the SVM (Hastie *et al*., 2009); in *fitcsvm*, the parameter is referred to as the “box constraint”. The reason for this name is that in the formulation of the dual optimization problem, the Lagrange multipliers are bounded to be within the range [0, *C*]. Hence, *C* poses a box constraint on the Lagrange multipliers.

In addition to the box constraint, *fitcsvm* routinely optimizes another hyperparameter that can impact the performance of the classifier: the kernel scale. All elements of the predictor matrix are divided by the kernel scale before the linear kernel norm is applied to compute the Gram matrix.

### Supplementary Material S6: BOLD activity during perception of emotional faces

Face-sensitive activity was assessed by means of a random effects group analysis (Friston *et al*., 1995) yielding brain regions that responded more strongly to faces than to scrambled images (*p*<0.05, family-wise error (FWE)-corrected at the peak level). In line with previous literature (Haxby *et al*., 2000), we found face-sensitive BOLD activity to be primarily located within the inferior occipital lobe and the fusiform gyrus in both hemispheres, referring to OFA and FFA, respectively (Supplementary Figure S3 and Supplementary Table S1). We also observed pronounced activity to faces in the left and right amygdala, a key region involved in emotion processing (e.g., Adolphs, 2002; Murray, 2007; Duvarci and Pare, 2014; Janak and Tye, 2015). Additionally, face-sensitive activation was observed in several other regions (see Supplementary Table S1), including the inferior frontal gyrus.

### Supplementary Material S7: Effective connectivity analysis

#### Sanity check: Accuracy of model fits

Effective connectivity among the regions of the extended face perception network was assessed using DCM for fMRI (Friston *et al*., 2003). We here focused on effective connectivity among OFA, FFA and amygdala (each in both hemispheres) given their well-established role as the core network mediating the perception of emotional faces (Haxby *et al*., 2000). Prior to harvesting the inferred connectivity parameters for group comparisons (see below) and classification analyses (see main text), we verified that the DCMs accounted for a reasonable amount of variance in the empirical fMRI data and that no group differences in the quality of model fit were present. To this end, we computed for each model in every participant the Pearson correlation between the predicted and measured time series of the entire BOLD signal – that is, collapsed across all regions of interest (Supplementary Figure S4A). The correlation coefficient ranged on average from *r*=0.32 (*p*<0.001) for model 7 (i.e., no modulation) to *r*=0.51 (*p*<0.001) for model 3 (i.e., modulation on all intra- and interhemispheric connections), which was the model with the highest negative free energy in most participants. Note that correlation coefficients were Fisher z-transformed to assess statistical significance. This suggests that the DCMs captured – at least to some degree – the characteristics of the empirical fMRI data and did not produce “flat-lines” without notable deviations of the posterior from the prior. Importantly, we did not find evidence for pronounced differences in the accuracy of the model fit across patient subgroups (one-way between-subject ANOVA with the factor *group*; all *F*_(2,82)_<0.82, *p*>0.45).

#### Effective connectivity patterns

Random-effects Bayesian model selection (Stephan *et al*., 2009; Rigoux *et al*., 2014) suggested model 3 to be the winning model at the group level with an expected posterior probability of 0.49 and a protected exceedance probability close to 1 (Supplementary Figure S4B). Nevertheless, at the single-subject level, other models received non-negligible posterior mass as well. To account for this variability, individual connectivity parameters were estimated using Bayesian model averaging (BMA; Penny *et al*., 2010) over all seven models in the model space within the default Occam’s window (*π*_*occ*_ = 0.05). Importantly, to avoid cross talk between training and test sample in the leave-one-out cross validation procedure (see Supplementary Material S2), average posterior densities were computed for each participant individually.

Individual BMA parameter estimates entered into summary statistics at the group level (one-sample t-tests, FDR-corrected for multiple comparisons). Supplementary Figure S5 summarizes the group-level effective connectivity patterns (averaged across all patient subgroups). In brief, we found excitatory driving inputs by the presentation of faces (regardless of the emotional valence) to the left and right OFA, consistent with its role as an early face-sensitive region in the brain (Puce *et al*., 1996; Rossion *et al*., 2012). Additionally, we observed excitatory intrahemispheric forward connections from OFA to FFA and from FFA to the amygdala, in both hemispheres. We also observed pronounced interhemispheric connectivity from left to right FFA, as well as from right to left FFA and from left to right OFA (although the latter two did not survive multiple comparisons correction).

With regard to modulatory influences by the emotional valence of the faces, we found several endogenous connections to show a significant modulation (see Supplementary Figure S5). In brief, modulatory effects of emotional valence were primarily observed for the feedforward intrahemispheric connections both between OFA and FFA, as well as between FFA and amygdala. This is consistent with previous work on effective connectivity within the face perception network (Fairhall and Ishai, 2007). In contrast, almost none of the feedback intrahemispheric connections was modulated by emotional valence; the only notable exception being the inhibitory modulation of the connection from right amygdala to right FFA by sad faces. Additionally, emotional valence modulated interhemispheric connections at the hierarchical level of the FFA (although this was significant only at an uncorrected threshold).

In summary, the effective connectivity patterns were rather consistent with previous findings of functional integration in the (extended) face perception network.

#### Group differences in effective connectivity

In a next step, we tested whether there are significant differences in the effective connectivity patterns across the different patient subgroups (two-sample t-tests). However, under appropriate multiple comparisons correction based on the false discovery rate (Benjamini and Yekutieli, 2001), no significant group differences were observed. At a more liberal threshold (*p*<0.05, uncorrected), some differences between the MDD subgroups were observed (see Supplementary Figure S6). In brief, effective connectivity patterns differed mainly in how emotions modulated functional integration among the face-processing (i.e., bilateral OFA and FFA) and emotion-sensitive regions (i.e., bilateral amygdala). More specifically, patients with a chronic disease trajectory differed from patients with gradual improvement of symptoms in the inhibitory self-connection of the left amygdala (*p*=0.005), in the modulatory influence of happy faces on the connection from right amygdala to right FFA (*p*=0.02), in the modulatory influence of angry faces on the connection from right to left amygdala (*p*=0.03) and left OFA to left FFA (*p*=0.04), in the modulatory influence of fearful faces on the connection from right FFA to right amygdala (*p*=0.01) and right OFA to right FFA (*p*=0.003), as well as in the modulatory influence of sad faces on the connection from left to right amygdala (*p*=0.02) and right FFA to right amygdala (*p*=0.01). Furthermore, patients with a chronic disease trajectory differed from patients showing fast remission in the inhibitory self-connection of the left amygdala (*p*=0.04), in how fearful faces modulated the connection from right FFA to right amygdala (*p*=0.03) and right OFA to right FFA (*p*=0.02), as well as in how sad faces modulated the connection from left FFA to left amygdala (*p*=0.047). Finally, patients with a gradual improvement in symptom severity differed from patients showing fast remission in the modulatory influence of happy faces on the connections from left amygdala to left FFA (*p*=0.049) and right FFA to right OFA (*p*=0.03), in the modulatory influence of fearful faces on the connections from right to left amygdala (*p*=0.03) and from right OFA to right FFA (*p*=0.02), as well as in the modulatory influence of sad faces on the connection from left to right amygdala (*p*=0.008).

While – as highlighted above – none of these group differences survived multiple comparisons correction, our findings suggest that some (small) alterations in effective connectivity exist between chronic, gradually improving and fast remitting patients, which might enable classification when jointly considering all DCM parameters as input features.

### Supplementary Material S8: Classification of clinical trajectories accounting for age

Since the three MDD groups (i.e., REM, IMP, and CHR) differed significantly in age (but no other variable; see Table 1 in the main text), we repeated the classification analysis when regressing out age as a confound from the DCM parameters.

Again, we found that DCM parameter estimates discriminate patients with a chronic disease trajectory from those showing fast remission with a balanced accuracy of 72% (*p*=0.001; Supplementary Figure S7, *blue*), corresponding to an area under the curve (AUC) of 0.83 (Supplementary Table S2). Furthermore, effective connectivity parameters discriminated between patients that showed gradual improvement of depressive symptoms and patients with fast remission with a balanced accuracy of 60% (*p*=0.04; Supplementary Figure S7). Again, this did not survive multiple comparisons correction (*α*_*Bonf*_=0.0056). The AUC was 0.66 (Supplementary Table S2). In contrast, chronic patients could not be differentiated from gradually improving patients above-chance level (balanced accuracy: 50%, *p*=0.63; Supplementary Figure S7), corresponding to an AUC of 0.28 (Supplementary Table S2).

As in the main text, we compared the GE results to measures of functional connectivity and local BOLD activity. First, we found that functional connectivity measures did not discriminate between any of the clinical trajectories above chance, with balanced accuracies of 49% for CHR vs. REM patients, 52% for CHR vs. IMP patients, and 49% for IMP vs. REM patients (all *p* > 0.05; Supplementary Figure S7, *light grey*). Similarly, estimates of local BOLD activity did not distinguish the different clinical trajectories, with balanced accuracies of 47% for CHR vs. REM patients, 47% for CHR vs. IMP patients, and 54% for IMP vs. REM patients (all *p* > 0.05; Supplementary Figure S7, *dark grey*).

Again, for discriminating patients with a chronic disease trajectory from those showing fast remission, GE significantly outperformed functional connectivity (*p*=0.02) and local BOLD activity measures (*p*=0.01).

In summary, the classification results obtained when accounting for age as a confounding variable are highly consistent to the ones reported in the main text – although balanced accuracies were slightly decreased. This suggests that classification was not simply caused by group difference in age.

## Figures

**Supplementary Figure S1:**
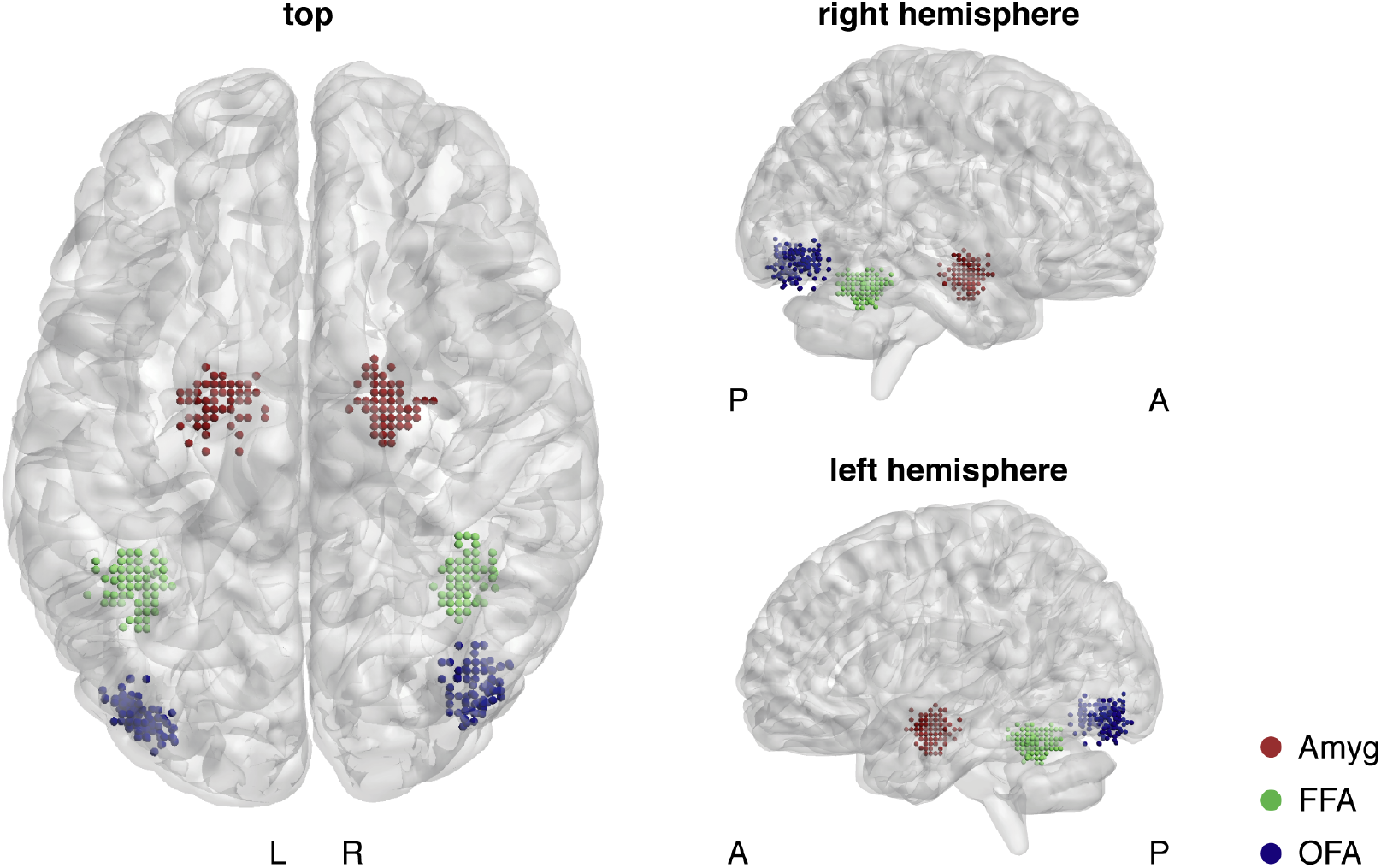
Individual locations of the regions of interest, given by the center coordinates of the spheres. Individual center coordinates were given as the nearest local maximum close to the MNI coordinates provided by Neurosynth (see Methods) with a 12mm sphere. Location of individual center coordinates were visualized using the BrainNet Viewer (Xia et al., 2013), publicly available for download (http://www.nitrc.org/projects/bnv/). Homotopic regions in the left and the right hemisphere are shown in the same color. L = left hemisphere; R = right hemisphere; A = anterior; P = posterior.

**Supplementary Figure S2:**
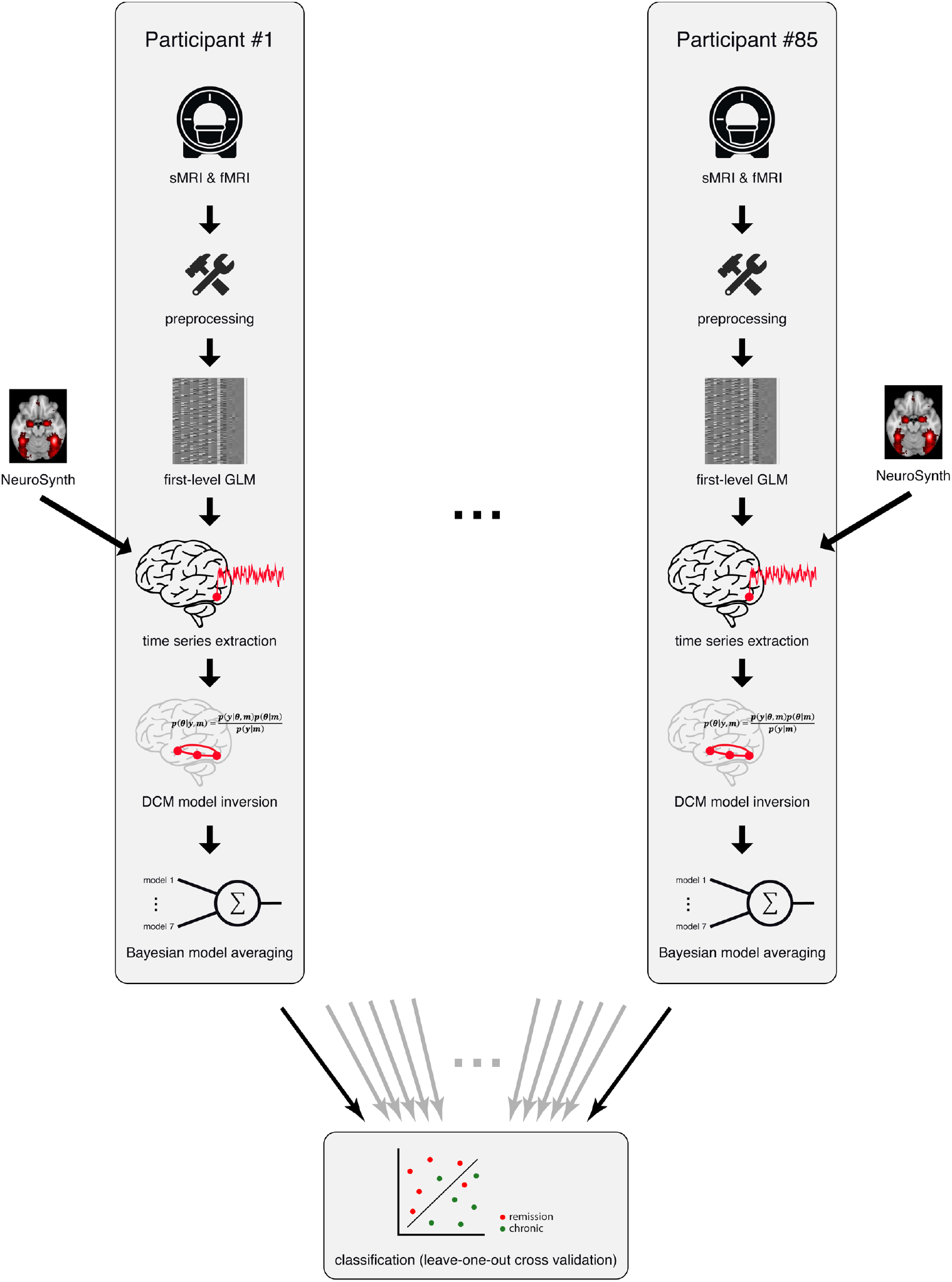
Schematic illustration of the generative embedding workflow utilized in the present study to ensure unbiased classification estimates. sMRI = structural magnetic resonance imaging; fMRI = functional magnetic resonance imaging; GLM = general linear model; DCM = dynamic causal model.

**Supplementary Figure S3:**
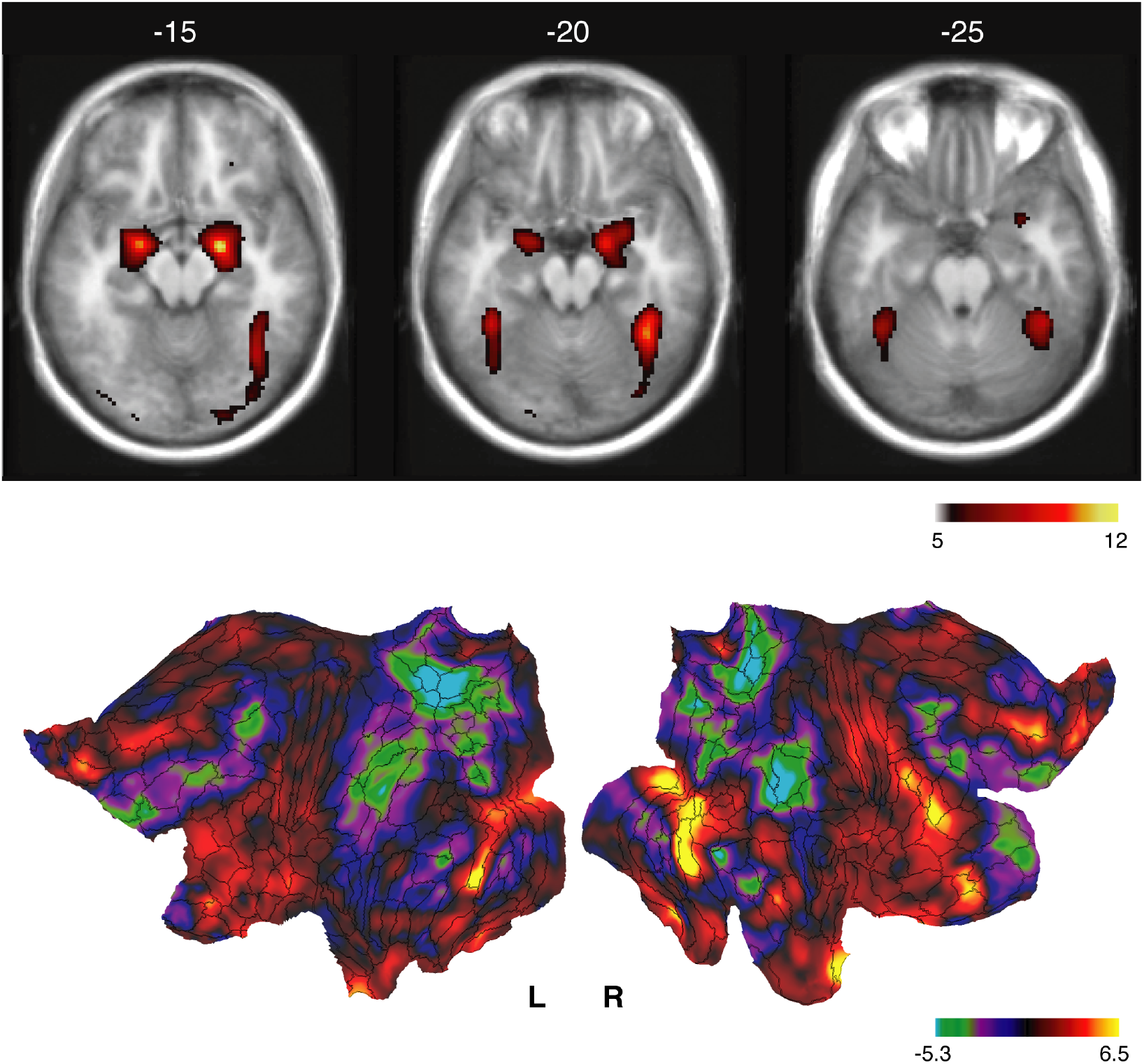
BOLD activity during perception of faces across all clinical groups (N=85). Activation pattern shows regions that were more activated during the perception of faces (regardless of the emotional valence) compared to scrambled faces. *Top:* Thresholded activation patterns were displayed on an anatomical template volume (*p*<0.05; family-wise error (FWE)-corrected at the peak level and a minimal cluster size of 5 voxels). *Bottom:* Unthresholded activation patterns displayed on a flat map of the cerebral cortex. Results were visualized using the Human Connectome Workbench (https://www.humanconnectome.org), and its predecessor Caret (http://brainvis.wustl.edu/wiki/index.php/Caret:About). L = left hemisphere, R = right hemisphere.

**Supplementary Figure S4:**
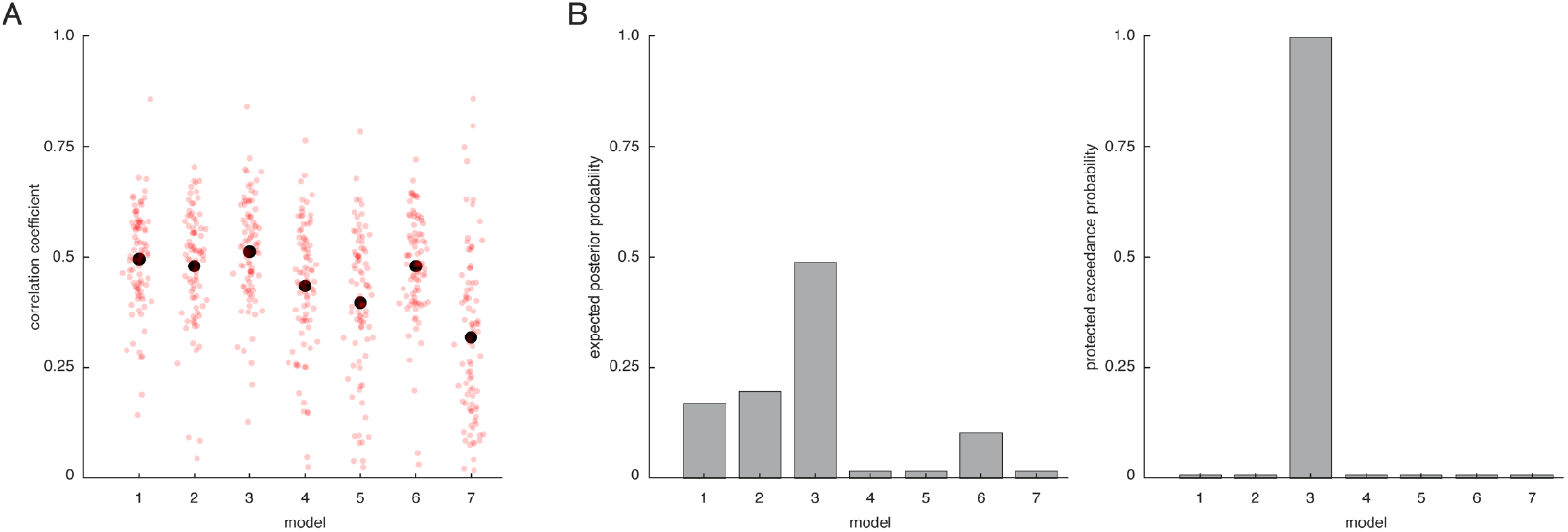
**(A)** Accuracy of model fit of the different DCMs for the face perception dataset from the NESDA study. Shown is the correlation between the measured and predicted time series across the entire BOLD signal (i.e., collapsed across all regions of interest). The small red circles represent the individual participants, whereas the black circle indicates the average across all participants. **(B)** Random-effects Bayesian model selection (BMS) results, illustrating (*left*) the expected posterior probability and (*right*) the protected exceedance probability.

**Supplementary Figure S5:**
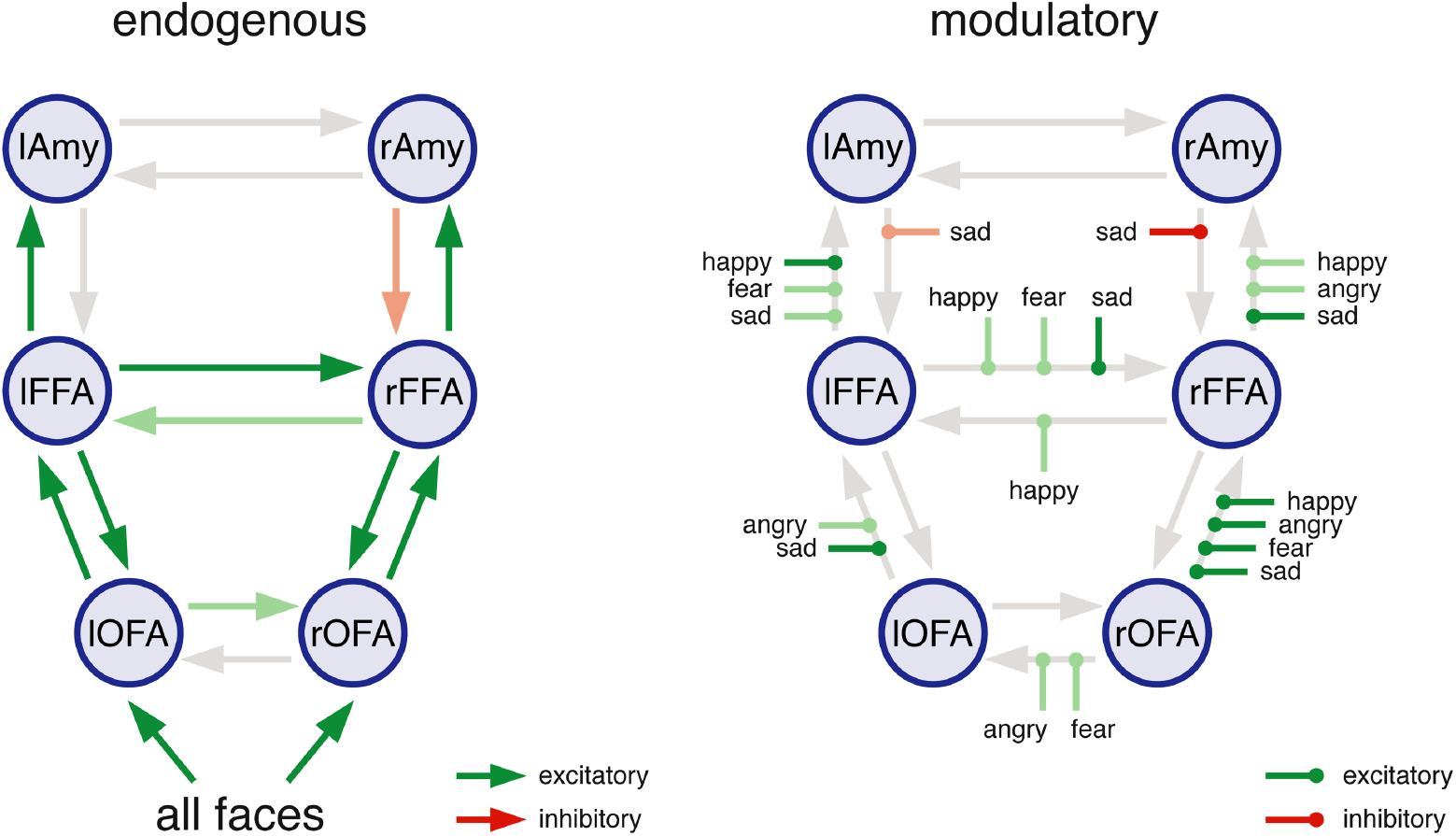
Group-level effective connectivity patterns (averaged across all patient subgroups). Significance of all group-level endogenous, modulatory and driving input parameters was assessed using one-sample t-tests based on the individual BMA parameter estimates of all 85 participants. Green arrows represent excitatory influences, whereas red arrows indicate inhibitory influences. Significant (FDR-corrected for multiple comparisons) parameters are shown in full color, parameters significant at an uncorrected threshold (*p*<0.05) are shown in faded colors. Furthermore, endogenous parameters that were not significant even at an uncorrected threshold are displayed in faded grey. lAmy = left amygdala, rAmy = right amygdala, lFFA = left fusiform face area, rFFA = right fusiform face area, lOFA = left occipital face area, rOFA = right occipital face area.

**Supplementary Figure S6:**
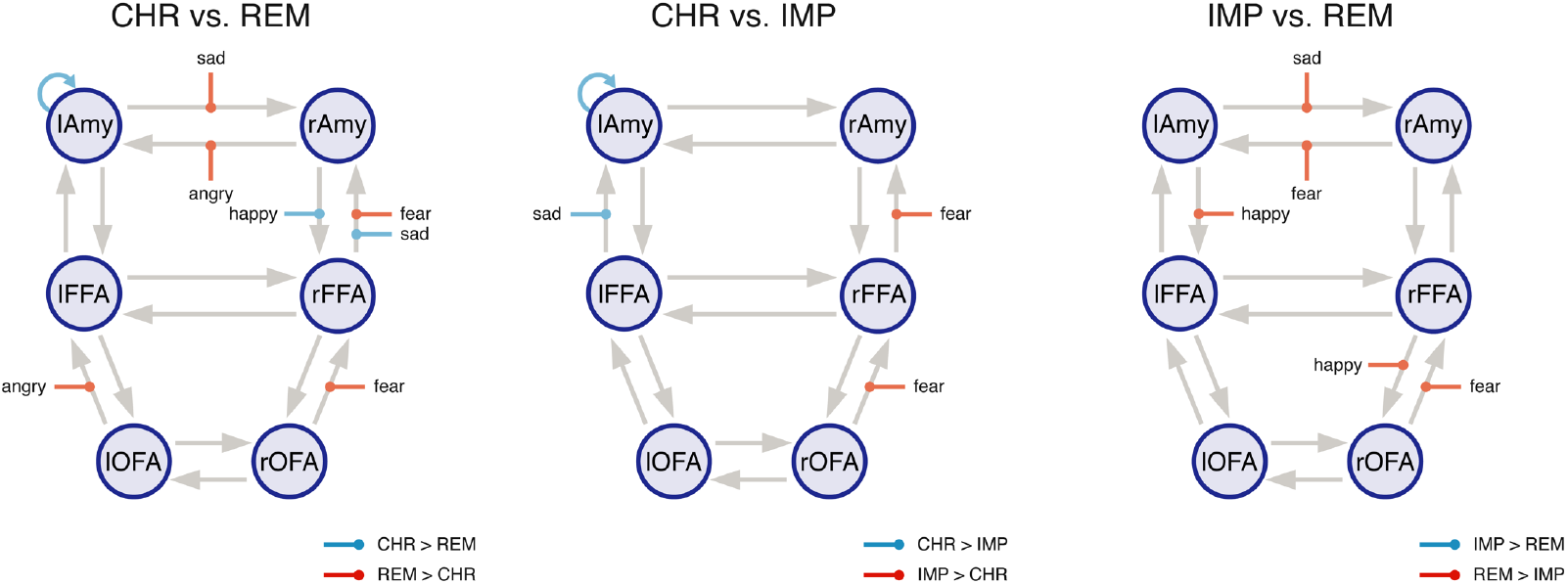
Differences in the effective connectivity patterns between the different MDD patient subgroups with distinct disease trajectories. Significance of group differences of all endogenous, modulatory and driving input parameters was assessed using two-sample t-tests. No significant group differences were observed when correcting for multiple comparisons (FDR-correction), but only at an uncorrected threshold (*p*<0.05). (*Left*) Blue arrows indicate parameters that were higher in CHR patients compared to REM patients. Red arrows indicate parameters that were higher in REM patients compared to CHR patients. (*Middle*) Blue arrows indicate parameters that were higher in CHR patients compared to IMP patients. Red arrows indicate parameters that were higher in IMP patients compared to CHR patients. (*Right*) Blue arrows indicate parameters that were higher in IMP patients compared to REM patients. Red arrows indicate parameters that were higher in REM patients compared to IMP patients. lAmy = left amygdala, rAmy = right amygdala, lFFA = left fusiform face area, rFFA = right fusiform face area, lOFA = left occipital face area, rOFA = right occipital face area, CHR = chronic disease trajectory, IMP = gradual improvement of symptom severity, REM = fast remission.

**Supplementary Figure S7:**
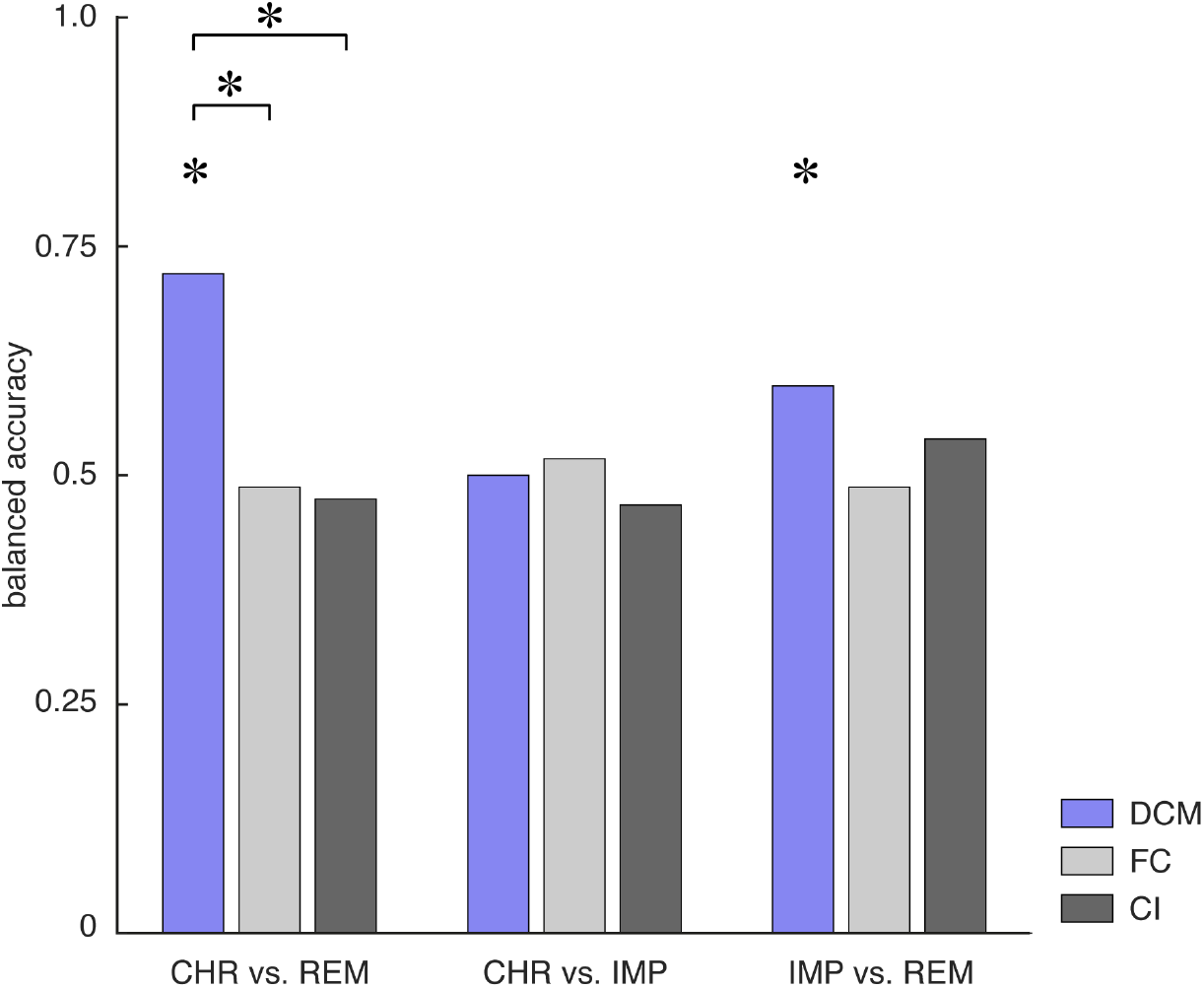
Balanced accuracy for the binary classifiers as assessed using leave-one-out cross validation for the three different subgroup comparisons – that is, CHR vs. REM (*left*), CHR vs. IMP (*middle*), and IMP vs. REM (*right*) – when accounting for age as a confound. Balanced accuracies are shown for the different features – namely, effective connectivity parameters (DCM; *blue*), functional connectivity (FC; *light grey*), and local BOLD activity (CI, *dark grey*). Asterisks above the bars indicate significant classification performance as assessed by means of permutation tests where an empirical null distribution of the balanced accuracy is computed by randomly permuting the participant labels and re-evaluating the classifier based on these new labels. Additionally, asterisks above the lines connecting two bars indicate significant differences in classification performance between different data features as assessed using the asymptotic McNemar test.

## Tables

**Supplementary Table S1:** MNI coordinates, cluster sizes and T scores for face-sensitivity activations (all faces > scrambled images). Shown are brain regions where activation is greater during the perception of faces (regardless of the emotional valence) as compared to the perception of scrambled images. Results are thresholded at a voxel-level threshold of *p*<0.05, FWE-corrected and a minimal cluster size of 5 voxels. Anatomical localization of the BOLD activations was performed using the Anatomy toolbox extension (Eickhoff *et al*., 2005).

| Cortical region | Hemisphere | MNI coordinates |  |  | Cluster size | T-score |
| --- | --- | --- | --- | --- | --- | --- |
|  |  | x | y | z |  |  |
| *Faces > scrambled* |  |  |  |  |  |  |
| Amygdala (**Amy**) | R | 20 | -8 | -16 | 642 | 11.69 |
| Amygdala (**Amy**) | L | -20 | -6 | -16 | 396 | 10.50 |
| Fusiform gyrus (**FFA**) | R | 40 | -52 | -20 | 1097 | 10.50 |
| Inferior occipital (**OFA**) | R | 46 | -80 | -8 |  | 7.70 |
| Fusiform gyrus (**FFA**) | L | -40 | -48 | -22 | 343 | 9.41 |
| Inferior frontal gyrus | R | 50 | 28 | 16 | 148 | 7.77 |
| Inferior frontal gyrus | R | 28 | 34 | -12 | 23 | 6.14 |
| Precentral gyrus | R | 44 | 2 | 30 | 34 | 6.01 |
| Inferior occipital (**OFA**) | L | -44 | -82 | -6 | 5 | 5.91 |
| Superior medial gyrus | R | 8 | 60 | 14 | 12 | 5.87 |
| Medial occipitotemporal gyrus | L | -22 | -94 | -18 | 6 | 5.65 |
| Inferior occipital | L | -38 | -84 | -16 | 11 | 5.56 |

**Supplementary Table S2:** Classification results for the generative embedding procedure when regressing out age as a confound from the DCM parameters. Shown are key performance measures of the classification algorithm, including: balanced accuracy, area under the curve, sensitivity (recall), specificity, positive predictive value (precision), and negative predictive value. Performance measures are shown for the three different binary classifications (i.e., CHR vs. REM, CHR vs. IMP, and IMP vs. REM).

| Classification | CHR ($n=15$)<br>vs. REM ($n=39$) | CHR ($n=15$)<br>vs. IMP ($n=31$) | IMP ($n=31$) vs.<br>REM ($n=39$) |
| --- | --- | --- | --- |
| Accuracy | 0.83 | 0.67 | 0.61 |
| Balanced accuracy | 0.72 | 0.50 | 0.60 |
| Area under the curve (AUC) | 0.83 | 0.28 | 0.66 |
| Sensitivity (recall) | 0.97 | 1.00 | 0.74 |
| Specificity | 0.47 | 0 | 0.45 |
| Positive predictive value (Precision) | 0.83 | 0.67 | 0.63 |
| Negative predictive value | 0.88 | 0 | 0.58 |

## Footnotes

1 The McNemar test is, strictly speaking, only valid when applied to a completely independent test set. Hence, comparing cross-validated classifiers might yield somewhat optimistic results. Having said this, for the present scenario, it is unclear what a statistically rigorous way would be to compare classifiers. For a discussion around this issue, see Dietterich (1998).

## Notes

### Competing Interest Statement

The authors have declared no competing interest.

### Author Declarations

All relevant ethical guidelines have been followed and any necessary IRB and/or ethics committee approvals have been obtained.

Any clinical trials involved have been registered with an ICMJE-approved registry such as ClinicalTrials.gov and the trial ID is included in the manuscript.

## REFERENCES

Ai H, Opmeer EM, Marsman JC, Veltman DJ, van der Wee NJA, Aleman A, et al. Longitudinal brain changes in MDD during emotional encoding: effects of presence and persistence of symptomatology. Psychol Med 2019: 1–11.

Almeida JR, Versace A, Mechelli A, Hassel S, Quevedo K, Kupfer DJ, et al. Abnormal amygdala-prefrontal effective connectivity to happy faces differentiates bipolar from major depression. Biol Psychiatry 2009; 66(5): 451–9.

Anand A, Li Y, Wang Y, Gardner K, Lowe MJ. Reciprocal effects of antidepressant treatment on activity and connectivity of the mood regulating circuit: an FMRI study. J Neuropsychiatry Clin Neurosci 2007; 19(3): 274–82.

Andrade L, Caraveo-Anduaga J, Berglund P, Bijl R, De Graaf R, Vollebergh W, et al. The epidemiology of major depressive episodes: results from the International Consortium of Psychiatric Epidemiology (ICPE) Surveys. International Journal of Methods in Psychiatric Research 2003; 12(1): 3–21.

Ashburner J, Friston KJ. Unified segmentation. Neuroimage 2005; 26(3): 839–51.

American Psychiatric Association. Diagnostic and Statistical Manual of Mental Disorders (DSM-5 R): American Psychiatric Publishing; 2013.

Bishop CM. Pattern recognition and machine learning: Springer, New York. 12, 13, 47, 105; 2006.

Breiter HC, Etcoff NL, Whalen PJ, Kennedy WA, Rauch SL, Buckner RL, et al. Response and habituation of the human amygdala during visual processing of facial expression. Neuron 1996; 17(5): 875–87.

Brodersen KH, Deserno L, Schlagenhauf F, Lin Z, Penny WD, Buhmann JM, et al. Dissecting psychiatric spectrum disorders by generative embedding. Neuroimage Clin 2014; 4: 98–111.

Brodersen KH, Schofield TM, Leff AP, Ong CS, Lomakina EI, Buhmann JM, et al. Generative embedding for model-based classification of fMRI data. PLoS Comput Biol 2011; 7(6): e1002079.

Buxton R, Wong E, Frank L. Dynamics of blood flow and oxygenation changes during brain activation: The balloon model. Magn Reson Med 1998; 39(6): 855–64.

Catani M, Thiebaut de Schotten M. A diffusion tensor imaging tractography atlas for virtual in vivo dissections. Cortex 2008; 44(8): 1105–32.

Cawley GC, Talbot NLC. On Over-fitting in Model Selection and Subsequent Selection Bias in Performance Evaluation. Journal of Machine Learning Research 2010; 11: 2079–107.

Clarke S, Miklossy J. Occipital cortex in man - Organization of callosal connections, related myeloarchitecture and cytoarchitecture, and putative boundaries of functional visual areas. J Comp Neurol 1990; 298(2): 188–214.

Crowther A, Smoski MJ, Minkel J, Moore T, Gibbs D, Petty C, et al. Resting-State Connectivity Predictors of Response to Psychotherapy in Major Depressive Disorder. Neuropsychopharmacology 2015; 40(7): 1659–73.

Cuthbert BN, Insel TR. Toward the future of psychiatric diagnosis: the seven pillars of RDoC. BMC Med 2013; 11: 126.

Daunizeau J, David O, Stephan K. Dynamic causal modelling: A critical review of the biophysical and statistical foundations. Neuroimage 2011; 58(2): 312–22.

de Graaf R, ten Have M, van Gool C, van Dorsselaer S. Prevalence of mental disorders and trends from 1996 to 2009. Results from the Netherlands Mental Health Survey and Incidence Study-2. Soc Psychiatry Psychiatr Epidemiol 2012; 47(2): 203–13.

Demenescu LR, Renken R, Kortekaas R, van Tol MJ, Marsman JB, van Buchem MA, et al. Neural correlates of perception of emotional facial expressions in out-patients with mild-to-moderate depression and anxiety. A multicenter fMRI study. Psychol Med 2011; 41(11): 2253–64.

Dietterich TG. Approximate Statistical Tests for Comparing Supervised Classification Learning Algorithms. Neural Comput 1998; 10(7): 1895–923.

Dinga R, Marquand AF, Veltman DJ, Beekman ATF, Schoevers RA, van Hemert AM, et al. Predicting the naturalistic course of depression from a wide range of clinical, psychological, and biological data: a machine learning approach. Transl Psychiatry 2018; 8(1): 241.

Dunlop BW, Rajendra JK, Craighead WE, Kelley ME, McGrath CL, Choi KS, et al. Functional Connectivity of the Subcallosal Cingulate Cortex And Differential Outcomes to Treatment With Cognitive-Behavioral Therapy or Antidepressant Medication for Major Depressive Disorder. Am J Psychiatry 2017; 174(6): 533–45.

Fairhall SL, Ishai A. Effective connectivity within the distributed cortical network for face perception. Cereb Cortex 2007; 17(10): 2400–6.

Fan RE, Chen PH, Lin CJ. Working set selection using second order information for training support vector machines. Journal of Machine Learning Research 2005; 6: 1889–918.

Frässle S, Lomakina EI, Kasper L, Manjaly ZM, Leff A, Pruessmann KP, et al. A generative model of whole-brain effective connectivity. Neuroimage 2018a; 179: 505–29.

Frässle S, Lomakina EI, Razi A, Friston KJ, Buhmann JM, Stephan KE. Regression DCM for fMRI. Neuroimage 2017; 155: 406-21.

Frässle S, Paulus FM, Krach S, Schweinberger SR, Stephan KE, Jansen A. Mechanisms of hemispheric lateralization: Asymmetric interhemispheric recruitment in the face perception network. Neuroimage 2016; 124(Pt A): 977–88.

Frässle S, Yao Y, Schobi D, Aponte EA, Heinzle J, Stephan KE. Generative models for clinical applications in computational psychiatry. Wiley Interdiscip Rev Cogn Sci 2018b; 9(3): e1460.

Frazier PI. A Tutorial on Bayesian Optimization. arXiv e-prints; 2018.

Friston K, Harrison L, Penny W. Dynamic causal modelling. Neuroimage 2003; 19(4): 1273–302.

Friston K, Holmes A, Poline J, Grasby P, Williams S, Frackowiak R, et al. Analysis of fMRI time-series revisited. Neuroimage 1995; 2(1): 45–53.

Friston K, Mattout J, Trujillo-Barreto N, Ashburner J, Penny W. Variational free energy and the Laplace approximation. Neuroimage 2007; 34(1): 220–34.

Friston KJ, Fletcher P, Josephs O, Holmes A, Rugg MD, Turner R. Event-related fMRI: characterizing differential responses. Neuroimage 1998; 7(1): 30–40.

Friston KJ, Mechelli A, Turner R, Price CJ. Nonlinear responses in fMRI: the Balloon model, Volterra kernels, and other hemodynamics. Neuroimage 2000; 12(4): 466–77.

Fu CH, Williams SC, Brammer MJ, Suckling J, Kim J, Cleare AJ, et al. Neural responses to happy facial expressions in major depression following antidepressant treatment. Am J Psychiatry 2007; 164(4): 599–607.

Fu CH, Williams SC, Cleare AJ, Brammer MJ, Walsh ND, Kim J, et al. Attenuation of the neural response to sad faces in major depression by antidepressant treatment: a prospective, event-related functional magnetic resonance imaging study. Arch Gen Psychiatry 2004; 61(9): 877–89.

Fu CHY, Mourao-Miranda J, Costafrecla SG, Khanna A, Marquand AF, Williams SCR, et al. Pattern classification of sad facial processing: Toward the development of neurobiological markers in depression. Biological Psychiatry 2008; 63(7): 656–62.

Godlewska BR, Norbury R, Selvaraj S, Cowen PJ, Harmer CJ. Short-term SSRI treatment normalises amygdala hyperactivity in depressed patients. Psychological Medicine 2012; 42(12): 2609–17.

Good PI. Permutation tests : a practical guide to resampling methods for testing hypotheses. 2nd ed. New York: Springer; 2000.

Greicius MD, Flores BH, Menon V, Glover GH, Solvason HB, Kenna H, et al. Resting-state functional connectivity in major depression: abnormally increased contributions from subgenual cingulate cortex and thalamus. Biol Psychiatry 2007; 62(5): 429–37.

Groenewold NA, Opmeer EM, de Jonge P, Aleman A, Costafreda SG. Emotional valence modulates brain functional abnormalities in depression: evidence from a meta-analysis of fMRI studies. Neurosci Biobehav Rev 2013; 37(2): 152–63.

Gueorguieva R, Mallinckrodt C, Krystal JH. Trajectories of depression severity in clinical trials of duloxetine: insights into antidepressant and placebo responses. Arch Gen Psychiatry 2011; 68(12): 1227–37.

Harmer CJ, Goodwin GM, Cowen PJ. Why do antidepressants take so long to work? A cognitive neuropsychological model of antidepressant drug action. Brit J Psychiat 2009; 195(2): 102–8.

Haufe S, Meinecke F, Gorgen K, Dahne S, Haynes JD, Blankertz B, et al. On the interpretation of weight vectors of linear models in multivariate neuroimaging. Neuroimage 2014; 87: 96–110.

Haxby J, Hoffman E, Gobbini M. The distributed human neural system for face perception. Trends Cogn Sci 2000; 4(6): 223–33.

Hirshfeld-Becker DR, Gabrieli JDE, Shapero BG, Biederman J, Whitfield-Gabrieli S, Chai XQJ. Intrinsic Functional Brain Connectivity Predicts Onset of Major Depression Disorder in Adolescence: A Pilot Study. Brain Connectivity 2019; 9(5): 388–98.

Hofer S, Frahm J. Topography of the human corpus callosum revisited - Comprehensive fiber tractography using diffusion tensor magnetic resonance imaging. Neuroimage 2006; 32(3): 989–94.

Horesh N, Klomek AB, Apter A. Stressful life events and major depressive disorders. Psychiatry Res 2008; 160(2): 192–9.

Itani S, Rossignol M, Lecron F, Fortemps P. Towards interpretable machine learning models for diagnosis aid: A case study on attention deficit/hyperactivity disorder. PLoS One 2019; 14(4): e0215720.

Kanwisher N, McDermott J, Chun M. The fusiform face area: A module in human extrastriate cortex specialized for face perception. J Neurosci 1997; 17(11): 4302–11.

Kapur S, Phillips AG, Insel TR. Why has it taken so long for biological psychiatry to develop clinical tests and what to do about it? Mol Psychiatry 2012; 17(12): 1174–9.

Kessler RC, van Loo HM, Wardenaar KJ, Bossarte RM, Brenner LA, Cai T, et al. Testing a machine-learning algorithm to predict the persistence and severity of major depressive disorder from baseline self-reports. Mol Psychiatry 2016; 21(10): 1366–71.

Kohler S, Chrysanthou S, Guhn A, Sterzer P. Differences between chronic and nonchronic depression: Systematic review and implications for treatment. Depress Anxiety 2019; 36(1): 18–30.

Kruijshaar ME, Barendregt J, Vos T, de Graaf R, Spijker J, Andrews G. Lifetime prevalence estimates of major depression: an indirect estimation method and a quantification of recall bias. Eur J Epidemiol 2005; 20(1): 103–11.

Lundqvist D, Flykt A, Öhman A. The Karolinska Directed Emotional Faces (KDEF). Stockholm: Department of Neurosciences Karolinska Hospital 1998.

Lyketsos C, Nestadt G, Cwi J, Heithoff K, Eaton W. The Life Chart Interview - A Standardized Method to Describe The Course of Psychopathology. International Journal of Methods in Psychiatric Research 1994; 4(3): 143–55.

MacQueen GM. Magnetic resonance imaging and prediction of outcome in patients with major depressive disorder. J Psychiatry Neurosci 2009; 34(5): 343–9.

Mayberg HS. Limbic-cortical dysregulation: a proposed model of depression. J Neuropsychiatry Clin Neurosci 1997; 9(3): 471–81.

Mayberg HS, Brannan SK, Mahurin RK, Jerabek PA, Brickman JS, Tekell JL, et al. Cingulate function in depression: a potential predictor of treatment response. Neuroreport 1997; 8(4): 1057–61.

McMahon FJ, Insel TR. Pharmacogenomics and personalized medicine in neuropsychiatry. Neuron 2012; 74(5): 773–6.

Muller VI, Cieslik EC, Serbanescu I, Laird AR, Fox PT, Eickhoff SB. Altered Brain Activity in Unipolar Depression Revisited: Meta-analyses of Neuroimaging Studies. JAMA Psychiatry 2017; 74(1): 47–55.

Murphy SE, Norbury R, O’Sullivan U, Cowen PJ, Harmer CJ. Effect of a single dose of citalopram on amygdala response to emotional faces. Br J Psychiatry 2009; 194(6): 535–40.

Musliner KL, Munk-Olsen T, Laursen TM, Eaton WW, Zandi PP, Mortensen PB. Heterogeneity in 10-Year Course Trajectories of Moderate to Severe Major Depressive Disorder: A Danish National Register-Based Study. JAMA Psychiatry 2016; 73(4): 346–53.

Muthen B, Asparouhov T, Hunter AM, Leuchter AF. Growth modeling with nonignorable dropout: alternative analyses of the STAR*D antidepressant trial. Psychol Methods 2011; 16(1): 17–33.

Nadeem MSA, Zucker JD, Hanczar B. Accuracy-Rejection Curves (ARCs) for Comparing Classification Methods with a Reject Option. Jmlr Worksh Conf Pro 2010; 8: 65–81.

Naselaris T, Kay KN, Nishimoto S, Gallant JL. Encoding and decoding in fMRI. Neuroimage 2011; 56(2): 400–10.

Nord CL, Halahakoon DC, Limbachya T, Charpentier C, Lally N, Walsh V, et al. Neural predictors of treatment response to brain stimulation and psychological therapy in depression: a double-blind randomized controlled trial. Neuropsychopharmacology 2019; 44(9): 1613–22.

Ojala M, Garriga GC. Permutation Tests for Studying Classifier Performance. Journal of Machine Learning Research 2010; 11: 1833-63.

Pan PM, Sato JR, Salum GA, Rohde LA, Gadelha A, Zugman A, et al. Ventral Striatum Functional Connectivity as a Predictor of Adolescent Depressive Disorder in a Longitudinal Community-Based Sample. American Journal of Psychiatry 2017; 174(11): 1112–9.

Penninx B, Nolen W, Lamers F, Zitman F, Smit J, Spinhoven P, et al. Two-year course of depressive and anxiety disorders: Results from the Netherlands Study of Depression and Anxiety (NESDA). Journal of Affective Disorders 2011; 133(1-2): 76–85.

Penninx BW, Beekman AT, Smit JH, Zitman FG, Nolen WA, Spinhoven P, et al. The Netherlands Study of Depression and Anxiety (NESDA): rationale, objectives and methods. Int J Methods Psychiatr Res 2008; 17(3): 121–40.

Penny W, Stephan K, Daunizeau J, Rosa M, Friston K, Schofield T, et al. Comparing Families of Dynamic Causal Models. PLoS Comput Biol 2010; 6(3).

Phillips ML, Chase HW, Sheline YI, Etkin A, Almeida JR, Deckersbach T, et al. Identifying predictors, moderators, and mediators of antidepressant response in major depressive disorder: neuroimaging approaches. Am J Psychiatry 2015; 172(2): 124–38.

Pitcher D, Walsh V, Duchaine B. The role of the occipital face area in the cortical face perception network. Experimental Brain Research 2011; 209(4): 481–93.

Puce A, Allison T, Asgari M, Gore J, McCarthy G. Differential sensitivity of human visual cortex to faces, letterstrings, and textures: A functional magnetic resonance imaging study. J Neurosci 1996; 16(16): 5205–15.

Raman S, Deserno L, Schlagenhauf F, Stephan KE. A hierarchical model for integrating unsupervised generative embedding and empirical Bayes. J Neurosci Methods 2016; 269: 6–20.

Rasmussen CE, Williams CKI. Gaussian Processes for Machine Learning. Adapt Comput Mach Le 2005: 1–247.

Rhebergen D, Lamers F, Spijker J, de Graaf R, Beekman AT, Penninx BW. Course trajectories of unipolar depressive disorders identified by latent class growth analysis. Psychol Med 2012; 42(7): 1383–96.

Rive MM, van Rooijen G, Veltman DJ, Phillips ML, Schene AH, Ruhe HG. Neural correlates of dysfunctional emotion regulation in major depressive disorder. A systematic review of neuroimaging studies. Neurosci Biobehav Rev 2013; 37(10 Pt 2): 2529–53.

Robertson B, Wang LH, Diaz MT, Aiello M, Gersing K, Beyer J, et al. Effect of bupropion extended release on negative emotion processing in major depressive disorder: A pilot functional magnetic resonance imaging study. J Clin Psychiat 2007; 68(2): 261–7.

Robins L, Wing J, Wittchen H, Helzer J, Babor T, Burke J, et al. The Composite International Diagnostic Interview - An epidemiological instrument suitable for use in conjunction with different diagnostic systems and in different cultures. Archives of General Psychiatry 1988; 45(12): 1069–77.

Rush A, Trivedi M, Wisniewski S, Nierenberg A, Stewart J, Warden D, et al. Acute and longer-term outcomes in depressed outpatients requiring one or several treatment steps: A STAR*D report. American Journal of Psychiatry 2006; 163(11): 1905–17.

Schmaal L, Marquand AF, Rhebergen D, van Tol MJ, Ruhé HG, van der Wee NJ, et al. Predicting the Naturalistic Course of Major Depressive Disorder Using Clinical and Multimodal Neuroimaging Information: A Multivariate Pattern Recognition Study. Biol Psychiatry 2015; 78(4): 278–86.

Shawe-Taylor J, Cristianini N. Kernel Methods for Pattern Analysis: cambridge University Press; 2004.

Sheline YI, Barch DM, Donnelly JM, Ollinger JM, Snyder AZ, Mintun MA. Increased amygdala response to masked emotional faces in depressed subjects resolves with antidepressant treatment: an fMRI study. Biol Psychiatry 2001; 50(9): 651–8.

Shen YD, Yao JS, Jiang XY, Zhang L, Xu LY, Feng R, et al. Sub-hubs of baseline functional brain networks are related to early improvement following two-week pharmacological therapy for major depressive disorder. Human Brain Mapping 2015; 36(8): 2915–27.

Siegle GJ, Thompson WK, Collier A, Berman SR, Feldmiller J, Thase ME, et al. Toward clinically useful neuroimaging in depression treatment: prognostic utility of subgenual cingulate activity for determining depression outcome in cognitive therapy across studies, scanners, and patient characteristics. Arch Gen Psychiatry 2012; 69(9): 913–24.

Smith SM, Jenkinson M, Woolrich MW, Beckmann CF, Behrens TEJ, Johansen-Berg H, et al. Advances in functional and structural MR image analysis and implementation as FSL. Neuroimage 2004; 23: S208–S19.

Stephan KE, Schlagenhauf F, Huys QJ, Raman S, Aponte EA, Brodersen KH, et al. Computational neuroimaging strategies for single patient predictions. Neuroimage 2017; 145(Pt B): 180–99.

Stephan KE, Weiskopf N, Drysdale PM, Robinson PA, Friston KJ. Comparing hemodynamic models with DCM. Neuroimage 2007; 38(3): 387–401.

Stone M. Cross-Validatory Choice and Assessment of Statistical Predictions. Journal of the Royal Statistical Society Series B-Statistical Methodology 1974; 36(2): 111–47.

Stuhrmann A, Dohm K, Kugel H, Zwanzger P, Redlich R, Grotegerd D, et al. Mood-congruent amygdala responses to subliminally presented facial expressions in major depression: associations with anhedonia. J Psychiatry Neurosci 2013; 38(4): 249–58.

Symmonds M, Moran CH, Leite MI, Buckley C, Irani SR, Stephan KE, et al. Ion channels in EEG: isolating channel dysfunction in NMDA receptor antibody encephalitis. Brain 2018; 141(6): 1691–702.

Uher R, Muthen B, Souery D, Mors O, Jaracz J, Placentino A, et al. Trajectories of change in depression severity during treatment with antidepressants. Psychol Med 2010; 40(8): 1367–77.

Van Essen DC, Newsome WT, Bixby JL. The pattern of interhemispheric connections and its relationship to extrastriate visual areas in the macaque monkey. J Neurosci 1982; 2(3): 265–83.

van Tol MJ, van der Wee NJ, van den Heuvel OA, Nielen MM, Demenescu LR, Aleman A, et al. Regional brain volume in depression and anxiety disorders. Arch Gen Psychiatry 2010; 67(10): 1002–11.

Velligan DI, Weiden PJ, Sajatovic M, Scott J, Carpenter D, Ross R, et al. Strategies for addressing adherence problems in patients with serious and persistent mental illness: recommendations from the expert consensus guidelines. J Psychiatr Pract 2010; 16(5): 306–24.

Vogelzangs N, Beekman AT, van Reedt Dortland AK, Schoevers RA, Giltay EJ, de Jonge P, et al. Inflammatory and metabolic dysregulation and the 2-year course of depressive disorders in antidepressant users. Neuropsychopharmacology 2014; 39(7): 1624–34.

Vreeburg SA, Hoogendijk WJ, DeRijk RH, van Dyck R, Smit JH, Zitman FG, et al. Salivary cortisol levels and the 2-year course of depressive and anxiety disorders. Psychoneuroendocrinology 2013; 38(9): 1494–502.

Walsh E, Carl H, Eisenlohr-Moul T, Minkel J, Crowther A, Moore T, et al. Attenuation of Frontostriatal Connectivity During Reward Processing Predicts Response to Psychotherapy in Major Depressive Disorder. Neuropsychopharmacology 2017; 42(4): 831–43.

Wang L, Hermens DF, Hickie IB, Lagopoulos J. A systematic review of resting-state functional-MRI studies in major depression. J Affect Disord 2012; 142(1-3): 6–12.

Wiecki TV, Antoniades CA, Stevenson A, Kennard C, Borowsky B, Owen G, et al. A Computational Cognitive Biomarker for Early-Stage Huntington’s Disease. PLoS One 2016; 11(2): e0148409.

Woo CW, Chang LJ, Lindquist MA, Wager TD. Building better biomarkers: brain models in translational neuroimaging. Nat Neurosci 2017; 20(3): 365–77.

Yao Y, Raman SS, Schiek M, Leff A, Frassle S, Stephan KE. Variational Bayesian inversion for hierarchical unsupervised generative embedding (HUGE). Neuroimage 2018; 179: 604–19.

Yarkoni T, Poldrack RA, Nichols TE, Van Essen DC, Wager TD. Large-scale automated synthesis of human functional neuroimaging data. Nat Methods 2011; 8(8): 665–70.

Zeki S. Interhemispheric connections of prestriate cortex in monkey. Brain Res 1970; 19(1): 63-&.

Zilles K, Clarke S. Architecture, connectivity and transmitter receptors of human extrastriate cortex. Comparison with non-human primates. Cerebral Cortex: Extrastriate Cortex in Primates: Plenum Press; 1997. p. 673–742.

## References

Adolphs R. Neural systems for recognizing emotion. Curr Opin Neurobiol 2002; 12(2): 169–77.

Benjamini Y, Yekutieli D. The control of the false discovery rate in multiple testing under dependency. Ann Stat 2001; 29(4): 1165–88.

Duvarci S, Pare D. Amygdala microcircuits controlling learned fear. Neuron 2014; 82(5): 966–80.

Eickhoff S, Stephan K, Mohlberg H, Grefkes C, Fink G, Amunts K, et al. A new SPM toolbox for combining probabilistic cytoarchitectonic maps and functional imaging data. Neuroimage 2005; 25(4): 1325–35.

Friston K, Harrison L, Penny W. Dynamic causal modelling. Neuroimage 2003; 19(4): 1273–302.

Hastie T, Tibshirani R, Friedman J. The Elements of Statistical Learning: Data mining, Inference, and Prediction: New York: Springer; 2009.

Janak PH, Tye KM. From circuits to behaviour in the amygdala. Nature 2015; 517(7534): 284–92.

Madigan D, Raftery A. Model selection and accounting for model uncertainty in graphical models using Occam’s window. JASA 1994; 89(428): 1535–46.

Murray EA. The amygdala, reward and emotion. Trends Cogn Sci 2007; 11(11): 489–97.

Pereira F, Mitchell T, Botvinick M. Machine learning classifiers and fMRI: a tutorial overview. Neuroimage 2009; 45(1 Suppl): S199–209.

Rigoux L, Stephan KE, Friston KJ, Daunizeau J. Bayesian model selection for group studies - revisited. Neuroimage 2014; 84: 971–85.

Rossion B, Hanseeuw B, Dricot L. Defining face perception areas in the human brain: A large-scale factorial fMRI face localizer analysis. Brain and Cognition 2012; 79(2): 138–57.

Stephan K, Penny W, Daunizeau J, Moran R, Friston K. Bayesian model selection for group studies. Neuroimage 2009; 46(4): 1004–17.

